# Exploring the clinical consequences and genetic aetiology of adult weight trajectories

**DOI:** 10.1101/2021.10.04.21264526

**Authors:** Jiayi Xu, Jessica S. Johnson, Eating Disorders Working Group of the Psychiatric Genomics Consortium, Andreas Birgegård, Jennifer Jordan, Martin A. Kennedy, Mikael Landén, Sarah L. Maguire, Nicholas G Martin, Preben Bo Mortensen, Liselotte V. Petersen, Laura M. Thornton, Cynthia M. Bulik, Laura M. Huckins

**Affiliations:** Pamela Sklar Division of Psychiatric Genomics, Icahn School of Medicine at Mount Sinai, New York, NY, USA; Department of Genetics and Genomic Sciences, Icahn School of Medicine at Mount Sinai, New York, NY, USA; Department of Medical Epidemiology and Biostatistics, Karolinska Institutet, Stockholm, Sweden; Department of Psychological Medicine, University of Otago, Christchurch, New Zealand; Canterbury District Health Board, Christchurch, New Zealand; Department of Pathology and Biomedical Science, University of Otago, Christchurch, New Zealand; Department of Psychiatry and Neurochemistry, Institute of Neuroscience and Physiology, The Sahlgrenska Academy at the University of Gothenburg, Gothenburg, Sweden; InsideOut Institute, Charles Perkins Centre, The University of Sydney, Camperdown, Sydney, NSW, Australia; Genetics & Computational Biology Department, QIMR Berghofer Medical Research Institute, Brisbane, Queensland, Australia; The Lundbeck Foundation Initiative for Integrative Psychiatric Research (iPSYCH), Aarhus, Denmark; National Centre for Register-Based Research, Aarhus BSS, Aarhus University, Aarhus, Denmark; Centre for Integrated Register-based Research (CIRRAU), Aarhus University, Aarhus, Denmark; Department of Psychiatry, University of North Carolina at Chapel Hill, Chapel Hill, NC, USA; Department of Nutrition, University of North Carolina at Chapel Hill, Chapel Hill, NC, USA; Department of Psychiatry, Icahn School of Medicine at Mount Sinai, New York, NY, USA; Icahn Institute for Genomics and Multiscale Biology, Icahn School of Medicine at Mount Sinai, New York, NY, USA; Seaver Autism Center for Research and Treatment, Icahn School of Medicine at Mount Sinai, New York, NY, USA; Mental Illness Research, Education and Clinical Centers, James J. Peters Department of Veterans Affairs Medical Center, Bronx, NY, USA

## Abstract

**Background:** Longitudinal weight trajectories may reflect individual health status. We examined the genetic aetiology and clinical consequences of adult weight trajectories in males and females leveraging genetic and phenotypic data in the electronic health records (EHR) of the Bio*Me*™ Biobank.

**Methods:** We constructed four longitudinal weight trajectories using annual EHR-recorded weights (stable weight, weight gain, weight loss, or weight cycle) (n=21,487). After validating the accuracy of the trajectories (n=100), we conducted a hypothesis-free phenome-wide association study (PheWAS), including sex-stratified PheWAS, to identify diseases associated with each weight trajectory. We then performed a hypothesis-driven polygenic risk score (PRS) analysis on these weight trajectories, focusing on anorexia nervosa (AN) and depression—both commonly associated with weight changes.

**Findings:** Weight trajectory classification was highly accurate (accuracy, sensitivity, and specificity > 97% for all four trajectories). Hypothesis-free PheWAS analyses identified a significant association between depression and weight cycle (OR=1.4, p≤7.7×10^−16^) after Bonferroni correction, but not with weight gain or loss. Compared to other weight trajectories, we also observed a significant association of osteoporosis-related phecodes with weight loss in females only (OR_female_=1.4, p_female_≤ 1.4×10^−7^, OR_male_=0.8, p_male_≥ 0.18). AN-PRS was positively associated with weight loss trajectory among individuals without eating disorder diagnoses (OR_top vs. bottom 10% PRS_=1.95, p=0.00035). Consistent effect direction was observed across three ancestry groups. The AN-PRS-weight loss association was not attenuated by obesity-PRS (OR_top vs. bottom 10% PRS_=1.94).

**Interpretation:** Adult weight trajectory is associated with disease both phenotypically and genetically. Our PheWAS reveals unique relationships between diseases and weight trajectory patterns, including the association of depression and weight cycle trajectory in both males and females, and osteoporosis-weight loss trajectory association in females only. In addition, our PRS analysis suggests that adults with higher AN genetic risk are more likely to have a weight loss trajectory, and this association may be independent of BMI/obesity-related genetic pathways.

**Funding:** Klarman Family Foundation, NIMH.

**Research in Context Panel:** *Evidence before this study:* We used PubMed and medRxiv to search for phenome-wide association studies (PheWAS) of BMI/weight that have been published and/or are currently in preprint. For the weight PheWAS, we used search terms: “(phewas[tiab] OR phenome wide[tiab]) AND (weight[tiab] OR BMI[tiab] OR body mass index[tiab])” on PubMed, and “phewas weight”, “phewas BMI”, “phewas body mass index”, “phenome weight”, “phenome BMI”, or “phenome body mass index” for abstract or title search on medRxiv (up to March 17, 2021). The literature search identified 45 studies in total. From title screening, 13 of the studies were further reviewed, and 5 studies were ultimately included as relevant evidence of PheWAS on weight or BMI. These five PheWAS included four studies of adult populations of European ancestry, and one study conducted in children (ALSPAC). The weight-related exposure variables used in these studies were genetic variants of the obesity-associated *FTO* gene, BMI-associated SNPs, BMI PRS, BMI value, and obesity status. Through using BMI/obesity-related exposures, these published PheWAS identified comorbidities associated with obesity, including type 2 diabetes, sleep apnea, hypertension, edema, liver disease, asthma, bronchitis, and earlier age of puberty in at least two of the PheWAS. The childhood PheWAS found positive associations of BMI PRS with multiple biomarkers, including leptin, C-reactive protein, IL6, triglyceride, very low-density lipoprotein, and a negative association with high density lipoprotein. One BMI PheWAS published in 2020 observed that hyperlipidemia and gastroesophageal reflux disease were only significantly associated with BMI on a phenotypic level, but not on a genetic level (e.g., BMI or obesity SNPs), likely due to the small genetic effect of single genetic variants. Regarding the impact of anorexia nervosa (AN) and depression genetic risk on weight trajectory, we searched “anorexia nervosa[title] AND (weight[title] OR BMI[title] OR body mass index[title]) AND (genetic[tiab])” or “depression[title] AND (weight[title] OR BMI[title] OR body mass index[title]) AND (genetic[tiab])” on PubMed, and “anorexia polygenic weight” or “anorexia polygenic BMI” or “anorexia polygenic body mass index” or “depression polygenic weight” or “depression BMI” or “depression body mass index” for abstract or title search on medRxiv (up to March 17, 2021). The literature search identified 36 studies in total, and 21 were further reviewed through the title screening. No studies were identified that examined the effect of depression genetic risk on BMI or weight, and only two were included as relevant evidence of AN genetic risk on BMI and weight. Of these two studies, one was cross-sectional in a small adult sample (age 18-59, n=380), and the other was longitudinal in a children/young adult population in the ALSPAC cohort (age 10-24, n=8,654). BMI PRS was found to be associated with lower BMI cross-sectionally, and with weight loss over time only in females.

*Added value of this study:* In this study, we create a novel inflection-point based method to classify longitudinal weight trajectory using weights recorded in the EHR in a hospital-based biobank (Mount Sinai Bio*Me*™ Biobank), with an accuracy of 98% or higher through our validation study (n=100). With this validated phenotype of weight pattern over time (i.e., weight trajectory), our PheWAS analysis afforded us the opportunity to examine comorbidity across the weight spectrum and across time. We identified 143 diseases associated with weight cycle (e.g., depression, anemias, renal failure),13 diseases positively associated with weight gain trajectory (e.g., obesity, obstructive sleep apnea, edema), and 36 with weight loss (e.g., protein-calorie malnutrition, gastrointestinal complication, end stage renal disease), after Bonferroni correction, using 5% as the cutoff for clinically relevant weight change. All diseases were negatively associated with a stable weight trajectory. Furthermore, we performed, to our knowledge, the first sex-stratified PheWAS related to weight trajectory, and identified eight sex-stratified associations with weight gain (e.g., obstructive sleep apnea), eight with weight loss (e.g., osteoporosis), and ten with weight cycle (e.g., vitamin B-complex deficiencies). On a genetic level, our study fills in the gap of the impact of AN genetic risk on longitudinal weight changes in the adult population. Unlike the finding in adolescents in the ALSPAC study, which found an AN-PRS-weight loss trajectory association only in females, we found an association of higher AN genetic risk with weight loss trajectory in both men and women, with consistent effect direction observed across individuals with European, African, and Hispanic ancestry in the Bio*Me*™ Biobank. Additionally, this association of AN genetics with weight loss was independent of the influence of obesity/BMI related genetic variants on weight.

*Implications of all the available evidence:* PheWAS is an excellent tool for exploring comorbidities associated across the weight spectrum. Our PheWAS findings identify diseases with different weight patterns (e.g., depression and weight cycle), which may reflect characteristics of these diseases, including age of onset, progression pattern, severity, and chronicity (e.g., the episodic nature of depression with the weight cycle pattern). In addition, our sex-stratified PheWAS implicates the important role of sex in weight regulation in the presence of disease. Certain sub-populations may be at greater risk of weight loss in some disease states (e.g., women with osteoporosis) and may need targeted treatment to address nutritional needs and to prevent further weight loss. Our study also suggests that people who have high AN genetic risk are at greater risk of displaying a weight loss trajectory during adulthood. However, given the limited amount of variation in the outcome of interest (e.g., weight loss) explained by the AN-PRS, the PRS may have to be jointly modeled with other risk factors to predict weight loss more accurately, or to identify subgroups at risk of weight loss. In addition, given our finding that the effect of AN genetics on weight loss was minimally affected by the obesity-related genetics, and the previously reported low genetic correlation of −0.22 between AN and obesity in the 2019 AN GWAS, this may indicate that AN- and obesity-related weight changes might have unique genetic underpinnings. Future studies that assess the pathway-specific genetic risk on weight pattern will further our understanding of the genetic architecture of longitudinal weight trajectory.

## Introduction

Weight trajectory in adults can be clinically important, potentially serving as an indicator of underlying health status. For example, a meta-analysis of thirty studies showed that older adults who had a non-stable weight trajectory (either weight gain, weight loss, or weight cycle) had a higher mortality rate compared to those with stable weight.^1^ Phenome-wide association studies (PheWAS), which have the capacity to examine associations of an exposure of interest across the full disease phenome, allow us to test associations between weight trajectory and health status in large-scale electronic health records (EHR), which contain longitudinal data for a large patient population, including frequent objective weight measures, taken as part of typical clinic visits. To date, weight-related PheWAS have used body mass index (BMI)-related genetic scores^2– 4^/variants^5^ or average BMI values^6^ as the exposure of interest, but have not yet explored the phenome-wide consequences of longitudinal weight trajectories. Therefore, in this study we leveraged the multiple weight measures as well as codes for disease diagnoses in the Bio*Me*™ Biobank^7^ to identify diseases associated with weight gain, weight loss, and/or weight cycle in a general patient population, stratified by sex. Gaining insights into the associations of weight trajectory with disease status could aid in monitoring disease progression and predicting prognosis.

Genetic studies have shown an association between higher polygenic risk of obesity and having a weight gain trajectory during both childhood and adulthood.^8,9^ However, especially in adults, much less is known regarding the low end of the weight spectrum; that is, whether a higher genetic risk for anorexia nervosa (AN) is associated with adult weight loss trajectory, even among individuals without any clinical diagnoses. Therefore, in this study, we tested whether AN-PRS predicted weight trajectories among adults in our biobank. In addition, we studied the impact of depression-related genetics on weight, given that weight changes are always seen in individuals with AN or depression.^10,11^

AN, characterized by extremely low body weight, has one of the highest mortality rates of all psychiatric disorders, besides substance abuse.^12,13^ Patients with AN experience significant weight loss and maintain a dangerously low body weight, which can be life-threatening.^10^ Although weight may be regained through nutritional rehabilitation and psychotherapy,^10^ full recovery is only achieved by about 30% of adult patients, and relapse is common.^14,15^ This cycle of remission-relapse may result in a weight cycle pattern in AN patients. Depression, one of the most common mental disorders, is a major contributor to global disease burden.^16^ It is associated with both loss of appetite and significant weight loss, or increased appetite and weight gain.^11,17^ On a population level, longitudinal studies have shown that depression during adolescence was associated with adult obesity, especially among females.^18,19^ In addition, adults with depression or mood disorders at baseline were at higher risk of weight gain in subsequent years.^20,21^

Both AN and depression have a genetic basis.^22,23^ The genome-wide SNP heritability (h_SNP_^2^) of AN and depression was estimated to be about 15% and 9%, respectively on a liability scale,^22–24^ suggesting an important role of genetics in the etiology of AN and depression. To date, the largest genome-wide association study (GWAS) for AN in the Psychiatric Genomics Consortium (PGC) identified eight genetic loci associated with AN. It also reported negative genetic correlations of AN with anthropometric and metabolic-related traits, such as obesity, BMI, waist-to-hip ratio, body fat, type 2 diabetes, and insulin.^22^ GWAS studies on depression, on the other hand, have found positive genetic correlation of depression with traits like obesity, BMI, waist-to-hip ratio, body fat, and triglycerides.^23,24^ Taken together, these findings indicate that genetic variation associated with AN or depression could also be associated with weight and metabolic regulation.

Only one previous study thus far has examined the association of AN-PRS with weight trajectory across childhood and young adulthood (aged 10-24).^25^ This study found an association between higher AN genetic risk and weight loss trajectory only in females. Less is known about the association of depression genetic score with weight. In our study, by leveraging the data in the Bio*Me*™ Biobank EHR, we investigated whether AN and depression-PRS are associated with longitudinal weight changes among adults (aged 25-85) without a clinical diagnosis of an eating disorder and/or depression, and whether this association differs by sex. Furthermore, in order to examine if the genetic finding is applicable to other populations, we also explored whether the PRS associations are seen across different ancestry groups (i.e., European, African, Hispanic).

## Methods

### Study Population

The Bio*Me*™ Biobank is a patient-based EHR-linked biorepository at the Mount Sinai Medical Center. The patient population within Bio*Me*™ reflects the racial and ethnic diversity (28% African American, 37% Hispanic Latino, 30% European American, and 5% other ancestry), as well as the community-level disease burden and health disparities in and around New York City.^26^ Participants consented to link their electronic health records, which contain information including disease diagnoses (ICD codes), laboratory results, medications, procedures, orders, and vital signs, to their genotype data.^27^ A total of 31,705 participants enrolled in the Bio*Me*™ Biobank with available genotype information were included in this study. Genotyping was performed using the Illumina Global Screening Array (**Supplementary Methods)**.^27,28^ The usage of data in this study complies with the regulation by the Health Insurance Portability and Accountability Act and is in accordance with the Mount Sinai Institutional Review Board standards.

### Weight Trajectory

Weight was measured at each physician visit. Annual weight was calculated as an average of weights in kilograms (kg) within a calendar year. All participants with at least 3 annual weight measures were included to construct the longitudinal weight trajectory. Extensive data cleaning was performed (detailed in **Supplementary Methods**), and the final weight dataset included 21,487 participants with 162,783 annual weight measures.

We created weight trajectories following two weight change thresholds: 5%^29^ and 10%^30^, based on previous evidence for clinical relevance. We generated four types of weight trajectories: stable weight, weight gain, weight loss, and weight cycle (5% or 10%, detailed definitions in **Supplementary Methods**). Stable weight and weight cycle are mutually exclusive; as are weight gain and weight loss. However, weight cycle can accompany weight gain or loss (e.g., one person could have weight gain + weight cycle pattern, weight loss + weight cycle pattern, or weight cycle only).

### Bio*Me*™ Diagnoses

Bio*Me*™ EHR data include 13,659 ICD-10 codes for 31,282 participants. Since ICD codes are used primarily for billing, rather than research purposes, we converted 13,659 ICD codes into 1,135 phecodes (**Supplementary Methods**). Phecodes combine ICD codes into disease groupings (for example, the AN phecode, 305.21, includes F50.00 – unspecified AN, F50.01 restricting type AN, and F50.02 binge eating/purging type AN).

### Phenome-Wide Association Study (PheWAS) of Weight Trajectories

We tested for associations between weight trajectories and phecodes using the R PheWAS package^31^, adjusting for genotype-derived sex and ancestry, age (baseline), age^2^, body mass index (BMI at baseline year), smoking status (ever/never tobacco user), alcohol use status (yes/no), and number of doctor visits. We included only phecodes with an effective size (N_eff_) >100 (**equation 1**) and at least 20 cases and 20 controls.

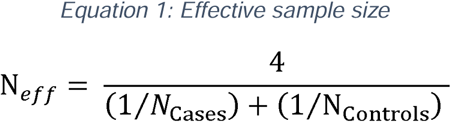

We established significance using a two-sided Bonferroni-corrected p-value of 4.4×10^−5^, accounting for 1,135 phecodes. Shared and distinct associations across and within each weight trajectory were examined through the UpSetR package in R (version 3.5.3).^32^ We also performed a sex-stratified PheWAS to discover sex-specific associations between disease and weight trajectories, with a nominal p-value of 0.05 to detect any difference in effect sizes between sex. In addition, we specifically reported any significant findings for neuropsychiatric disorders.

### Heritability and Genetic Correlation of Weight Trajectories

Weight trajectory heritabilities were estimated with GCTA-GREML using variants with minor allele frequency (MAF) > 0.0001.^33^ To estimate the heritability of non-stable weight trajectories (i.e., weight gain, loss, cycle), we included only individuals with stable weight trajectories as controls, whereas to estimate the heritability of stable weight trajectory, we included all other trajectories as controls. Genetic correlations among four weight trajectories were estimated using bivariate GREML analysis,^34^ adjusted for baseline BMI. The bias induced by sample overlap between each pairs of weight trajectories is minimal via GREML.^35^

### Construction of PRS in Bio*Me*™

We calculated AN and MDD-PRS in the Bio*Me*™ population using the most recent PGC GWAS for both disorders.^22,24^ We also calculated obesity-PRS as a positive control for weight gain trajectory.^36^ Obesity PRS was selected instead of BMI PRS so as to avoid the sample overlap between the base and target samples for PRS calculation, as Bio*Me*™ participants are included in several recent BMI GWAS.^37,38^ We used summary statistics of obesity class I (BMI ∈ [30,34.9]), rather than class II (BMI ∈ [35,39.9]) or III (BMI ≥ 40), as a positive control, given its larger sample size (32,858 cases, 2.6-fold greater sample size). PRS were calculated for individuals without any ICD-10 codes for eating disorder (ED, F50) and depression (F32 and F33), given our main goal to investigate the genetic risk of AN and depression on weight change in individuals without clinical diagnoses. Details on PRS calculation and quality control (QC) are in **Supplementary Methods**.

Since these base GWAS included participants of predominantly European ancestry, our primary PRS analyses were performed in Bio*Me*™ BioBank participants of European ancestry (N= 4,979). We performed 10,000 permutations to reduce the type I error (i.e., false positives) by generating an empirical p-value for each PRS-weight trajectory association. Exploratory analyses were performed in other ancestries (African American and Hispanic Latino) to further examine the external validity of any suggestive association of AN- or depression-PRS with weight trajectories found in the European ancestry population. For the purposes of this analysis, we classified individuals according to both self-report and genotype-derived ancestry. The three ancestry groups (European, African, Hispanic) are mutually exclusive to each other in our analysis (i.e., no overlap of individuals across these three groups) based on their genotype and self-report of race and ethnicity. Individuals with discordant self-report and genotype-derived ancestry definitions were excluded from this analysis.

### Adjustment of Covariates

We adjusted for age (baseline), age^2^, BMI (kg/m^2^, at baseline), sex, disease/health history that may influence weight (i.e., cancer, COPD, HIV, hypothyroidism, end stage renal disease, bariatric surgery, and pregnancy, with details on how these were coded in **Supplementary Methods**), and genotype-derived principal components (PC) 1-5 to account for population stratification in our PRS analysis. Sensitivity analyses were performed to adjust for smoking status and alcohol use status.

For the depression-PRS, we performed sensitivity analyses adjusting for the use of commonly prescribed antidepressants to test whether any depression-weight trajectory relationship is mediated through side-effects of these medications. The antidepressants selected as covariates included different drug categories, such as tricyclic antidepressants (amitriptyline, amoxapine, desipramine, doxepin, imipramine, nortriptyline, protriptyline), monoamine oxidase inhibitors (phenelzine, tranylcypromine), selective serotonin reuptake inhibitors (citalopram, escitalopram, fluoxetine, paroxetine, sertraline), serotonin-noradrenaline/norepinephrine reuptake inhibitors (duloxetine, venlafaxine), and others (mirtazapine, trazodone).

### Statistical analysis

Follow-up PRS analyses that examined the association of PRS deciles (e.g., top versus bottom decile) with weight trajectories (e.g., weight loss trajectory vs. stable weight as the reference group) were performed in R (version 3.5.3). Sensitivity analyses of sex-modification of the association of PRS with weight trajectories were also carried out by including an interaction term of PRS and sex for any of the significant findings from the main PRS analysis. In order to compare the genetic and phenotypic associations of AN and depression with weight trajectories, we also tested the association of eating disorder (305.2) and depression (296.2) phecodes with the weight trajectories in the European ancestry participants. We used the eating disorder phecode (305.2, n=9) rather than AN (305.21) since very few European ancestry individuals with AN had all covariate information available (n=1). Further conditional analyses were performed to assess the independence of AN-PRS, depression-PRS, and obesity-PRS with regard to their effects on longitudinal weight trajectory.

## Results

### Accurate phenotyping of weight trajectories in BioMe™

A total of 20,550 participants aged 25 to 85 were included in this study: 61.8% of these participants were female (**Table 1**), and 32.5% of the participants fell in the overweight category (BMI ∈ [25,29.9]). On average, each individual had seven annual weights (range 3-18) across a nine-year span in their EHR (range 3-22 years). The largest weight changes per year per individual ranged from −59 kg to +30 kg.

**Table 1.**
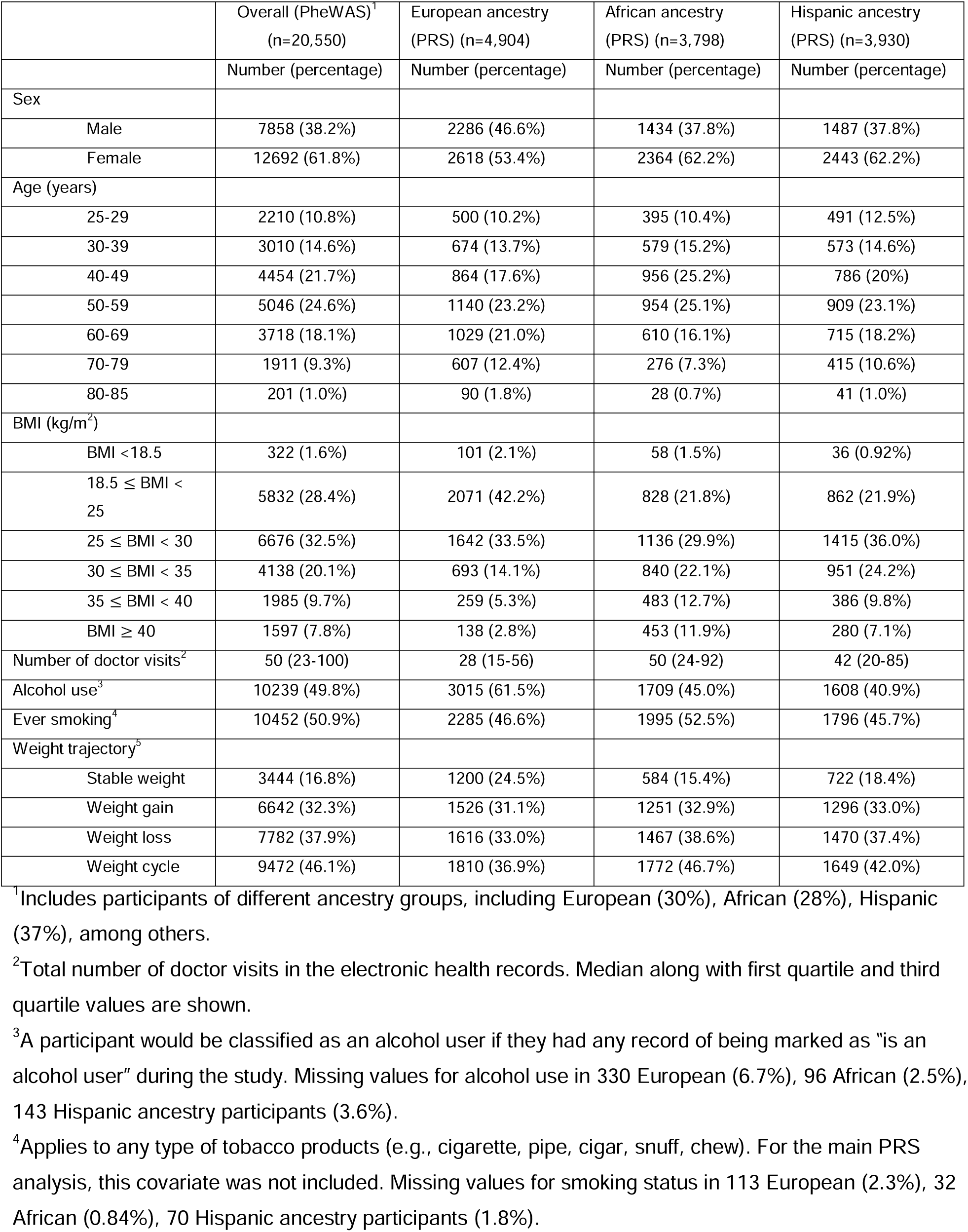
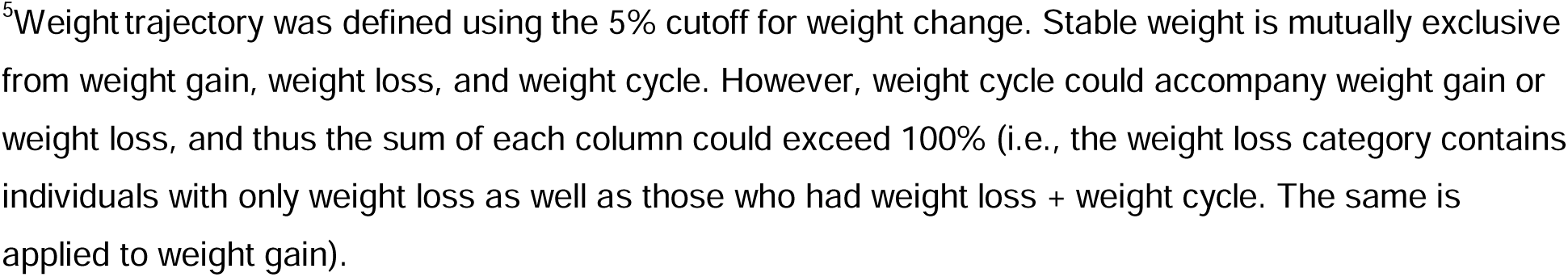
Characteristics of participants included in PheWAS and PRS analyses.

|  | Overall (PheWAS)<sup>1</sup><br>(n=20,550) | European ancestry<br>(PRS) (n=4,904) | African ancestry<br>(PRS) (n=3,798) | Hispanic ancestry<br>(PRS) (n=3,930) |
| --- | --- | --- | --- | --- |
|  | Number (percentage) | Number (percentage) | Number (percentage) | Number (percentage) |
| Sex |  |  |  |  |
| Male | 7858 (38.2%) | 2286 (46.6%) | 1434 (37.8%) | 1487 (37.8%) |
| Female | 12692 (61.8%) | 2618 (53.4%) | 2364 (62.2%) | 2443 (62.2%) |
| Age (years) |  |  |  |  |
| 25-29 | 2210 (10.8%) | 500 (10.2%) | 395 (10.4%) | 491 (12.5%) |
| 30-39 | 3010 (14.6%) | 674 (13.7%) | 579 (15.2%) | 573 (14.6%) |
| 40-49 | 4454 (21.7%) | 864 (17.6%) | 956 (25.2%) | 786 (20%) |
| 50-59 | 5046 (24.6%) | 1140 (23.2%) | 954 (25.1%) | 909 (23.1%) |
| 60-69 | 3718 (18.1%) | 1029 (21.0%) | 610 (16.1%) | 715 (18.2%) |
| 70-79 | 1911 (9.3%) | 607 (12.4%) | 276 (7.3%) | 415 (10.6%) |
| 80-85 | 201 (1.0%) | 90 (1.8%) | 28 (0.7%) | 41 (1.0%) |
| BMI (kg/m<sup>2</sup>) |  |  |  |  |
| BMI <18.5 | 322 (1.6%) | 101 (2.1%) | 58 (1.5%) | 36 (0.92%) |
| 18.5 ≤ BMI < 25 | 5832 (28.4%) | 2071 (42.2%) | 828 (21.8%) | 862 (21.9%) |
| 25 ≤ BMI < 30 | 6676 (32.5%) | 1642 (33.5%) | 1136 (29.9%) | 1415 (36.0%) |
| 30 ≤ BMI < 35 | 4138 (20.1%) | 693 (14.1%) | 840 (22.1%) | 951 (24.2%) |
| 35 ≤ BMI < 40 | 1985 (9.7%) | 259 (5.3%) | 483 (12.7%) | 386 (9.8%) |
| BMI ≥ 40 | 1597 (7.8%) | 138 (2.8%) | 453 (11.9%) | 280 (7.1%) |
| Number of doctor visits<sup>2</sup> | 50 (23-100) | 28 (15-56) | 50 (24-92) | 42 (20-85) |
| Alcohol use<sup>3</sup> | 10239 (49.8%) | 3015 (61.5%) | 1709 (45.0%) | 1608 (40.9%) |
| Ever smoking<sup>4</sup> | 10452 (50.9%) | 2285 (46.6%) | 1995 (52.5%) | 1796 (45.7%) |
| Weight trajectory<sup>5</sup> |  |  |  |  |
| Stable weight | 3444 (16.8%) | 1200 (24.5%) | 584 (15.4%) | 722 (18.4%) |
| Weight gain | 6642 (32.3%) | 1526 (31.1%) | 1251 (32.9%) | 1296 (33.0%) |
| Weight loss | 7782 (37.9%) | 1616 (33.0%) | 1467 (38.6%) | 1470 (37.4%) |
| Weight cycle | 9472 (46.1%) | 1810 (36.9%) | 1772 (46.7%) | 1649 (42.0%) |

<sup>1</sup>Includes participants of different ancestry groups, including European (30%), African (28%), Hispanic (37%), among others.
<sup>2</sup>Total number of doctor visits in the electronic health records. Median along with first quartile and third quartile values are shown.
<sup>3</sup>A participant would be classified as an alcohol user if they had any record of being marked as “is an alcohol user” during the study. Missing values for alcohol use in 330 European (6.7%), 96 African (2.5%), 143 Hispanic ancestry participants (3.6%).
<sup>4</sup>Applies to any type of tobacco products (e.g., cigarette, pipe, cigar, snuff, chew). For the main PRS analysis, this covariate was not included. Missing values for smoking status in 113 European (2.3%), 32 African (0.84%), 70 Hispanic ancestry participants (1.8%).
<sup>5</sup>Weight trajectory was defined using the 5% cutoff for weight change. Stable weight is mutually exclusive from weight gain, weight loss, and weight cycle. However, weight cycle could accompany weight gain or weight loss, and thus the sum of each column could exceed 100% (i.e., the weight loss category contains individuals with only weight loss as well as those who had weight loss + weight cycle. The same is applied to weight gain).

We created four weight trajectories: stable weight, weight gain, weight loss, and weight cycle (**S. Figure 1**). We assessed the sensitivity and specificity of our weight trajectory classifications through manual review of 100 randomly selected individuals (**S. Table 1-3**). Our random sample had similar weight trajectory distributions as in the overall sample (**S. Table 2**). Excellent sensitivity (≥ 97.2%), specificity (≥ 98.0%), and accuracy (≥ 98%) were achieved for the weight trajectory classifications (**S. Table 1**).

Using the 5% cutoff for weight change, only 16.8% of participants maintained a stable weight. Among those with a non-stable weight trajectory (n = 17,106), 15.7% had a weight cycle trajectory without any sustained weight gain or loss (n = 2,682), 16.8% had a weight cycle + weight gain trajectory (n = 2,873), 22.9% had a weight cycle + weight loss trajectory (n = 3,917), 22.0% had weight gain only (n = 3,769), and 22.6% had weight loss only (n = 3,865). Weight loss trajectory had a strongly negative correlation with weight gain trajectory both phenotypically and genetically (phenotypic correlation: r = −0.53, genetic correlation: r_g_ = −0.85), but a weak correlation with stable weight (phenotypic correlation: r = −0.36, genetic correlation: r_g_ = −0.23). Weight cycle was weakly correlated with weight loss or weight gain only on a genetic level with little phenotypic correlations (phenotypic level: r= 0.05 and −0.03, genetic level: r_g_ = 0.22 and −0.31, respectively) (**S. Figure 2**). Individuals who had a stable weight trajectory had fewest weight records in EHR (mean n = 5.7), followed by weight gain (n = 7.5) and weight loss (n = 7.9), whereas those with weight cycle had the most weight records in EHR on average (mean = 8.9, all pairwise p<5.1×10^−13^). In addition, baseline BMI distributions also varied across different weight trajectories such that individuals with weight loss trajectory had the highest BMI on average (mean BMI = 31.1, interquartile range (IQR) = [25.8, 34.8]), followed by those with weight cycle (mean BMI = 29.8, IQR = [24.6, 33.6]), stable weight (mean BMI = 27.9, IQR = [23.5, 30.9]), and weight gain (mean BMI = 27.5, IQR = [23.1, 30.6]).

Next, we derived SNP heritability estimates for all four weight trajectories. Stable weight, weight gain, weight loss, and weight cycle trajectories had heritabilities of 2.1%, 4.1%, 5.5%, and 4.7%, respectively (all p< 0.05, **S. Table 4**). Heritability was much greater when the weight change cutoff was set at 10% (stable weight: 9.1%, weight gain: 14.2%, weight loss: 19.0%, and weight cycle: 24.2%). In addition, heritability remained significant after adjusting for baseline BMI (**S. Table 4**).

### Weight trajectory PheWAS and sex-stratified PheWAS: depression, osteoporosis and more

We performed a PheWAS to identify traits associated with each of our weight trajectories using 5% as the cutoff for clinically relevant weight changes (see **Supplementary Results** for findings using the 10% cutoff). All phecodes that were significantly associated with stable weight had negative associations (**Figure 1D**), indicating a consistently negative relationship between disease and maintaining a stable weight. For example, depression was negatively associated with a stable weight trajectory (OR=0.63, p=7.5×10^−12^, **S. Table 5**).

**Figure 1.**
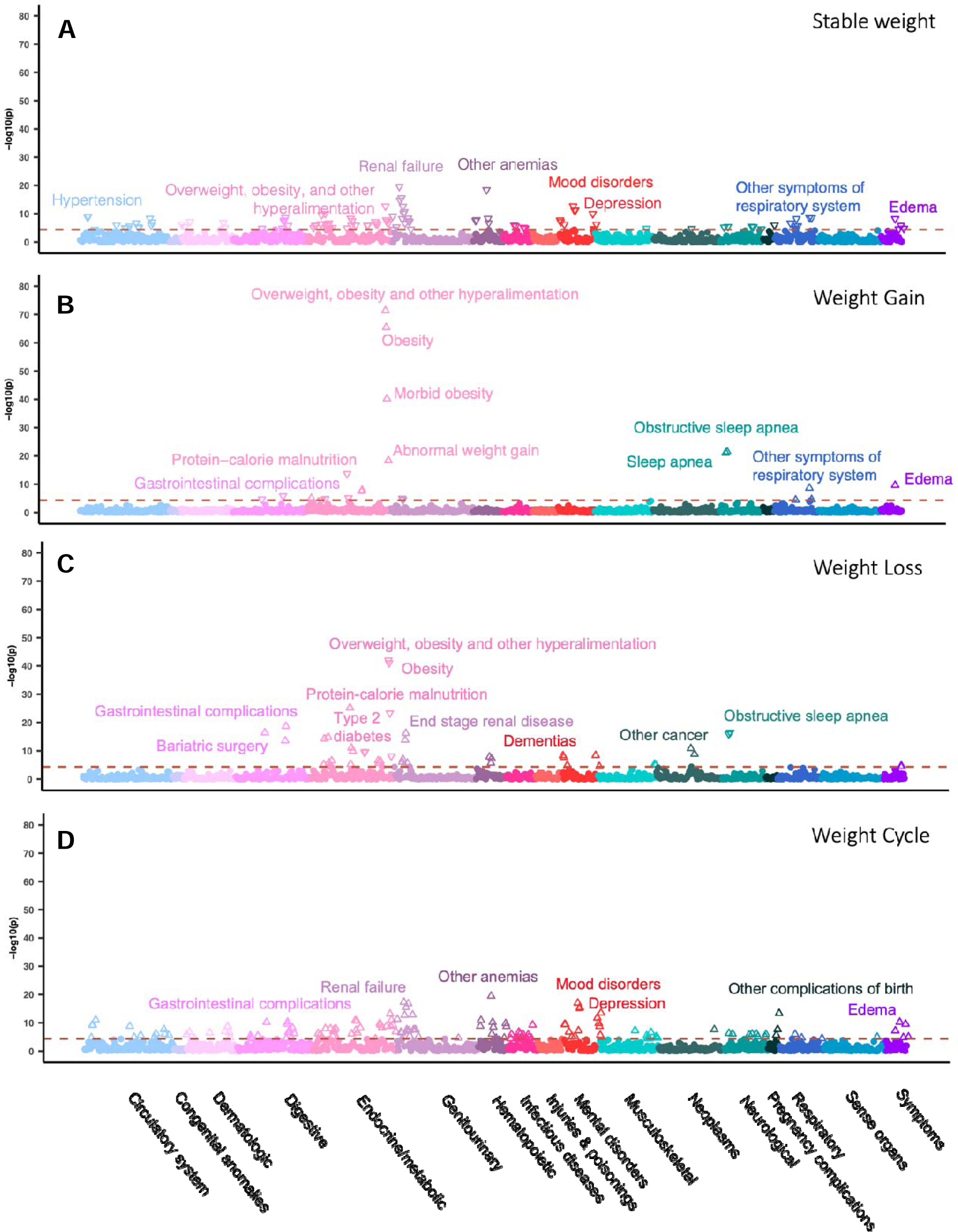
Phenome-wide association plots of weight trajectory with Bio*Me*™ Biobank phecodes using the 5% weight change cutoff. (A) PheWAS plot for stable weight trajectory; (B) PheWAS plot for weight gain trajectory; (C) PheWAS plot for weight loss trajectory; (D) PheWAS plot for weight cycle trajectory. Phecodes above the blue line passed the Bonferroni-corrected p-value threshold (P < 4.4×10^−5^). Phecodes are grouped into 17 different disease categories. An upward triangle (Δ) denotes a positive association, while a downward triangle (⍰) denotes a negative association. The top associations are annotated in each plot.

The clinical diagnoses demonstrating the strongest positive association with the weight gain trajectory were a cluster of obesity-related phecodes [278, 278.1, 278.11] (p ≤ 6·5×10^−41^; **Figure 1, S. Table 5**). In addition, we identified a number of respiratory outcomes associated with weight gain, including sleep apnea, shortness of breath, asthma with exacerbation, and other symptoms of the respiratory system (p ≤ 3.52×10^−5^; **S. Table 5**). The positive association of obstructive sleep apnea with weight gain trajectory was significantly greater in females than males (Odds Ratio (OR)_female_=2.2, p=4.1×10^−18^ *vs*. OR_male_=1.6, p=2.1×10^−6^, p_diff_=0.023, **S. Table 7, S. Figure 3**).

The phecodes with the strongest positive association with the weight loss trajectory were protein-calorie malnutrition (p=6.3×10^−26^), followed by gastrointestinal complications (p=2.1×10^−19^), bariatric surgery (p=4.0×10^−17^), and end stage renal disease (p=6.1×10^−17^) (**Figure 1, S. Table 5**). Diagnoses that are typically associated with weight loss were also significant in our PheWAS, including type 2 diabetes, cancer, anorexia, cachexia, substance addiction, and tobacco use disorder (p ≤ 3.42×10^−5^, **S. Table 5-6**). In addition, type 2 diabetes was significantly associated with weight loss, but not with weight gain or weight cycle (**S. Table 9-10**). Our sex-stratified analysis showed that osteoporosis-related phecodes (743, 743.1, 743.11) were significantly associated with weight loss in females (OR=1.4, p ≤ 1.4×10^−7^), but not in males (OR=0.8, p ≥ 0.18, **S. Table 7, S. Figure 4**), whereas anemia was significantly associated with weight loss in males, but not in females at a Bonferroni-corrected significance level (OR_male_=1.5, p=7.8×10^−10^, OR_female_=1.2, p=0.0044, p_diff_=0.0026, **S. Table 7-8**).

A total of 110 phecodes were uniquely associated with weight cycle trajectory, (i.e., reached Bonferroni-significance only for weight cycle), including depression-related phecodes (296.2, 296.22) (p ≤ 7.7×10^−16^, **S. Table 9-10**), hypertensive complications (p=8.9×10^−12^), and vitamin deficiency (p ≤ 1.6×10^−11^). Other significant phecodes included bipolar disorder, blood diseases, chronic liver disease, and viral hepatitis C (**S. Table 9**). Swelling of a limb and vitamin B complex deficiency were found to be more strongly associated with weight cycle in males (OR=2.1 and 1.8, respectively) than in females (OR=1.4 and 1.3, p_diff_=0.040 and 0.049, respectively, **S. Table 7, S. Figure 5**).

Several phecodes were associated with both weight cycle and weight loss trajectories, while some others were associated with both weight cycle and weight gain trajectories (**S. Table 9-10**). In particular, renal failure, anemia, bariatric surgery, and gastrointestinal complications (**S. Table 9-10**) were associated with both weight cycle and weight loss. Contrary to protein-calorie malnutrition and end stage renal disease (ESRD), which had greater effect magnitudes with weight loss than weight cycle (malnutrition: OR_weight loss_=4.3 and OR_weight cycle_=1.9, p_diff_=2.9×10^−5^, ESRD: OR_weight loss_=2.0 and OR_weight cycle_=1.5, p_diff_=0.029), renal failure tended to have a greater magnitude with weight cycle (OR=1.49) compared with weight loss (OR=1.25, p_diff_=0.0067, **S. Table 9**). Meanwhile, traits associated with both weight cycle and weight gain trajectories included morbid obesity, abnormal weight gain, and edema. Of these, morbid obesity and abnormal weight gain had stronger associations with the weight gain trajectory (OR=4.0 and 3.4, respectively) than the weight cycle trajectory (OR=1.9 and 2.3, p_diff_=1.1×10^−7^ and 0.047, respectively, **S. Table 9**), whereas the association strength of edema with weight gain and weight cycle trajectories was similar (OR=1.6 and 1.5, p_diff_=0.78). No disease was simultaneously positively associated with both weight gain and weight loss (**S. Figure 6**).

We further examined the associations of psychiatry- and neurology-related phecodes with weight trajectories. No phecodes in this category were associated with weight gain. Phecodes that were significantly associated with the weight cycle trajectory, but not the weight loss trajectory, included depression-related phecodes (296.2, 296.22), bipolar disorder, and alcoholic liver damage. Phecodes associated with both the weight cycle and weight loss trajectories included substance use disorder, tobacco use disorder, neurological disorders, and dementia (**S. Table 11**).

### Polygenic risk analysis: AN-PRS with weight loss trajectory

We tested whether our weight trajectories had shared genetic aetiology with psychiatric traits. First, as a positive control, we confirmed that obesity-PRS was significantly associated with the weight gain trajectory (empirical p=0.042; **S. Table 12**), and that individuals in the top obesity-PRS decile had increased likelihood of weight gain trajectory compared to the bottom decile (OR=1.38 using 5% weight gain cutoff; **Figure 2**).

**Figure 2.**
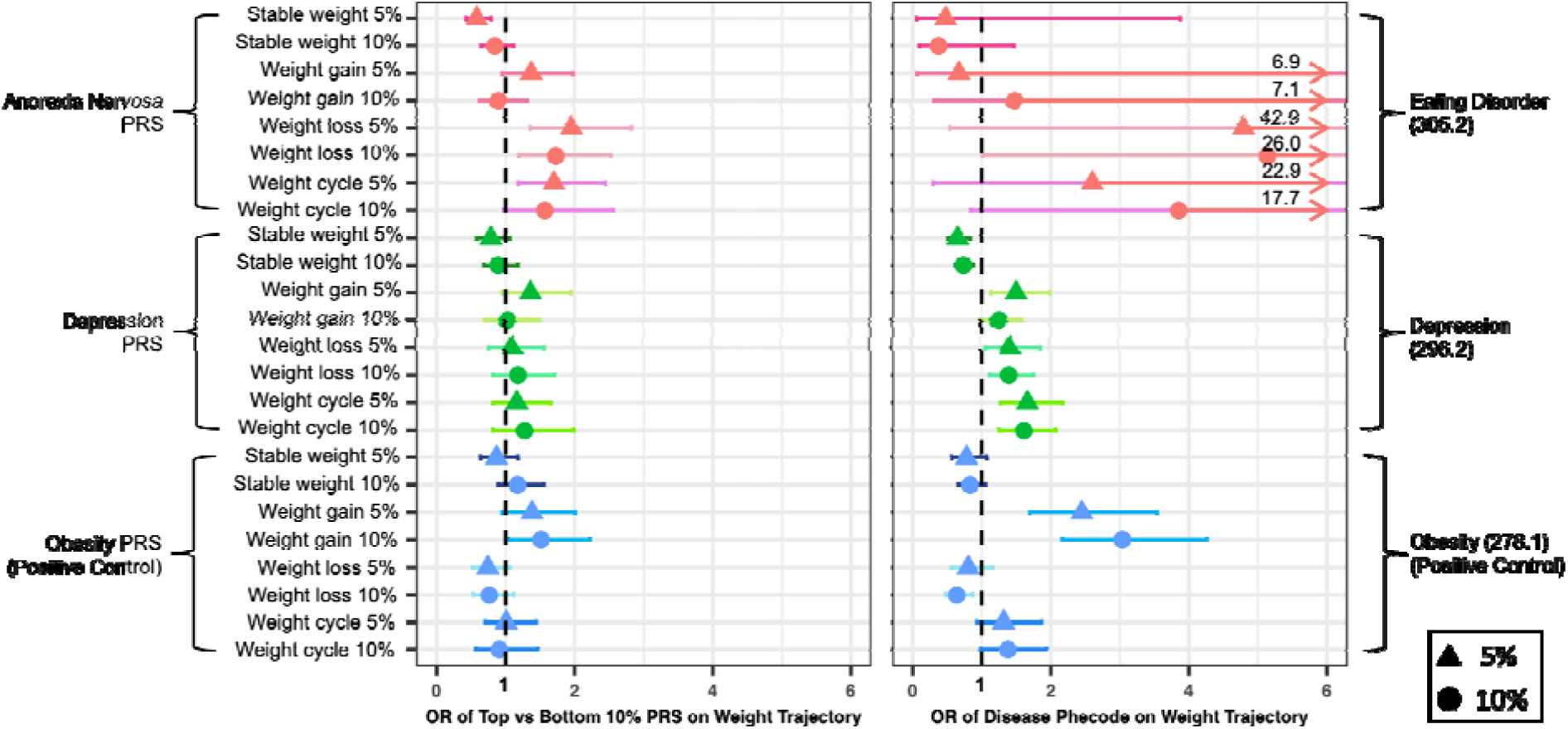
Association of AN and depression with weight trajectory in the European ancestry participants in the Bio*Me*™ Biobank. The left panel shows the odds ratio of individuals in top versus bottom decile of PRS (AN, depression, and obesity class I) with different weight trajectories (defined by either 5% or 10% weight change cutoff). The right panel shows the association of AN and depression with different weight trajectories on a phenotypic level using phecodes as the exposure. Individuals with stable weight was used as controls for those with weight gain, weight loss, and weight cycle trajectories. Given the small sample size of AN case in the European ancestry samples (n=1), its parent phecode, eating disorder, was used for the phenotypic association (n=9). Obesity-PRS and phecode are used as positive controls. Abbreviation: AN, anorexia nervosa; PRS, polygenic risk score.

Next, we tested the associations of AN-PRS with weight trajectories. We found that AN-PRS was positively associated with weight loss trajectory (empirical p=0.011, R^2^=0.52%, **S. Table 13**) and negatively associated with stable weight trajectory (empirical p=0.027, R^2^=0.29%, **S. Table 13**). Individuals in the top decile of AN-PRS risk (who did not have a clinical diagnosis of eating disorder (ICD-10 code of F50 in the EHR)) were twice as likely to have a weight loss trajectory (OR=1.95), and half as likely to have a stable weight trajectory (OR=0.58) compared to individuals in the bottom decile (**Figure 2-3, Table 2**). Besides using stable weight as the reference group for the AN-PRS-weight loss trajectory association, we also tested the association using everyone who did not have a weight loss trajectory as the comparison group (i.e., including those with weight gain or weight cycles) and found a consistent effect direction (top vs bottom PRS decile OR=1.42), though not significant (empirical p=0.097). On average, the AN-PRS scores were highest in individuals with weight loss trajectories, followed by stable weight, and then weight gain (**S. Figure 7**). We observed no sex modification effect (p=0.58) on the AN-PRS-weight loss trajectory association and no effect difference in those who had a weight loss trajectory with or without weight cycle (p=0.22). We also computed the AN-PRS in African and Hispanic ancestry groups using the base GWAS (PGC-ED) derived from European ancestry individuals. We observed consistent directions of effect among African American (OR top vs. bottom decile = 1.9) and Hispanic individuals (OR = 1.3), even though these estimates were not significant (p=0.27 and 0.63, respectively).

**Table 2.** Sensitivity analysis of the association of anorexia nervosa polygenic risk score (PRS) with weight loss trajectory.

| Model | OR<sub>top versus bottom decile</sub> | 95% CI | P |
| --- | --- | --- | --- |
| Original | 1.95 | [1.36, 2.82] | 0.00035 |
| Conditioning on obesity-PRS | 1.94 | [1.35, 2.81] | 0.00040 |
| Conditioning on depression-PRS | 1.92 | [1.33, 2.79] | 0.00050 |
| Additional adjustment of smoking status and alcohol use | 1.79 | [1.23, 2.62] | 0.0026 |

**Figure 3.**
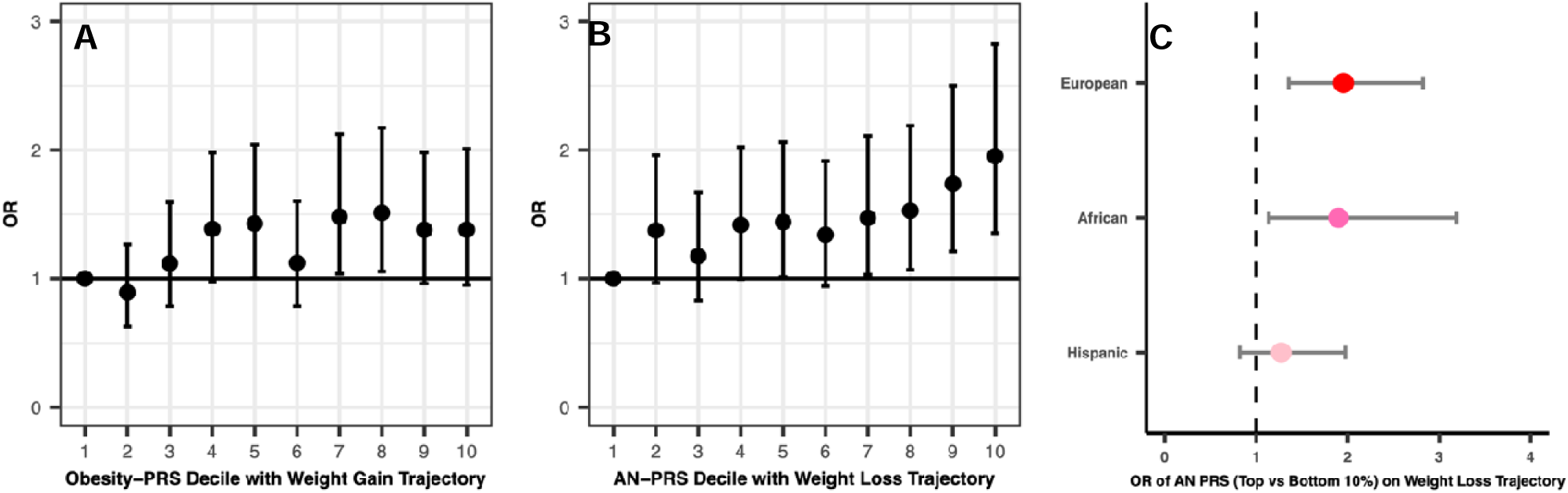
Associations of PRS with weight trajectory by deciles and by ancestry. (A) Odds ratio of each obesity-PRS decile with weight gain trajectory, using the bottom decile as the reference group. (B) Odds ratio of each AN-PRS decile with weight loss trajectory, using the bottom decile as the reference group. (C) The odds ratio of individuals in the top versus bottom decile of AN-PRS with weight loss trajectory (defined by 5% cutoff) across different ancestry groups (European, African, Hispanic). Abbreviation: AN, anorexia nervosa; PRS, polygenic risk score.

We performed further sensitivity analyses to test the mitigating effect of obesity PRS on the effect of AN-PRS on weight loss trajectory (**Table 2**). The AN-PRS-weight loss trajectory association changed minimally regardless of adjusting obesity-PRS or depression-PRS in the model. This indicated little genetic overlap between AN-PRS and obesity-PRS or depression-PRS with regards to their effects on weight loss trajectory.

This was confirmed by the minimal overlap between number of SNPs included for the AN, depression, and obesity-PRS (<1% overlap, **S. Figure 8**). The positive AN-PRS-weight loss association remained after further adjustment for lifestyle factors, including smoking and alcohol use status (more sensitivity analyses included in **Supplementary Results**).

Finally, we tested whether depression-PRS was significantly associated with weight trajectories, given the substantial evidence for shared genetic aetiology between depression and BMI,^23^ as well as evidence for the association of depression with weight trajectories in our PheWAS. However, depression-PRS was not associated with any of the weight trajectories (i.e., gain, loss, cycle) in our sample (empirical P ≥ 0.17, **S. Table 14**). Individuals in the top decile of depression-PRS risk did not have a significantly greater odds of having a weight gain, weight loss, or weight cycle trajectory, compared to those in the bottom decile of depression-PRS risk (OR=1.4 with 95% CI = [0.9,1.9]; OR=1.1 with 95% CI = [0.8,1.6]); OR=1.2 with 95% CI = [0.8,1.7], respectively). Furthermore, the depression-PRS-weight trajectory associations changed minimally after adjusting for antidepressant usage (OR=1.4, 95% CI = [0.9,1.9] with weight gain, OR=1.1, 95% CI = [0.8,1.5] with weight loss, OR=1.2, 95% CI = [0.8,1.7] with weight cycle).

## Discussion

Longitudinal weight changes can be an important indicator of individual health status over time. However, the association of longitudinal weight trajectory with the full disease spectrum, as well as its genetic aetiology, remain unclear. Our study identified four distinct longitudinal weight trajectories among adults and demonstrated that stable weight trajectory is associated with lower risk of disease. Individuals with a stable weight trajectory also have the fewest doctor visits in general as they have the fewest number of annual weight records in EHR compared to those with non-stable weight trajectories. Since our study concerns weight change, rather than absolute weight or BMI, we correct for BMI at baseline. Our association between stable weight and lower risk of disease, then, is striking; having a stable weight is associated with lower risk of disease, regardless of BMI. It is important, however, to keep in mind that having a stable weight could be a consequence of being disease-free, and it does not suggest that maintaining a stable weight at the extreme ends of the weight spectrum is health-promoting (e.g., morbidly underweight or obese), given the majority of people with a stable weight in our study had BMIs ranged from 21.1 (10th percentile) to 35.9 (90th percentile).

By contrast, we found strong associations between weight cycling and depression (OR=1.42, p=7.7×10^−16^, **S. Table 5**), in line with well-established roles for appetite and weight change as vegetative signs of depression. We also identified associations between bipolar disorder, substance use, and tobacco use with weight cycle and weight loss. Weight gain trajectories were further associated with an increased risk of respiratory disorders. Our PheWAS also identified sex-specific associations between weight trajectories and disease; for example, we found significantly higher associations between osteoporosis and weight loss among females vs. males (OR=1.4 vs 0.8). Although we cannot infer causality due to the cross-sectional nature of our study, this association is in line with previous evidence of sex disparities in osteoporosis.^39^ Future studies that examine the role of sex in weight regulation for different diseases and health conditions would be of interest.^40^

To date, no study has explored the genetic basis of longitudinal weight trajectories in adulthood. Here, we demonstrated that weight trajectory is significantly genetically heritable, even when adjusting for baseline BMI (**S. Table 4)**; that is, weight change is itself heritable and genetically regulated, regardless of underlying body weight. Interestingly, we observed a higher heritability of weight trajectory at the 10% cutoff than 5% cutoff, which could indicate a stronger genetic basis for individuals who are more susceptible to weight changes (e.g., some individuals may be prone to gain or lose more weight, or have bigger weight fluctuation, whereas some individuals could be more resistant to weight changes).

Given the roles of weight loss in AN aetiopathology and BMI genetics in AN risk, we next sought to establish whether polygenic risk for AN regulates weight loss among adults. Indeed, we found that higher AN-PRS was associated with a higher likelihood of weight loss trajectory. Our study is the first to demonstrate that AN-PRS regulates weight change among adults without clinical diagnoses of eating disorders. The use of obesity as a positive control, the dose-response relationship observed across AN-PRS deciles, the consistent effect direction across three ancestry groups (EA, AA, and HA), and the reduction of false positive rate through permutation strengthen our finding of the positive association of a higher AN-PRS with having a weight loss trajectory. We excluded any individuals with eating disorder or depression ICD-10 codes in the PRS analysis, so that the AN-PRS on weight is less likely to be driven by the disease itself and more likely to be driven by genetics. However, it is still possible that there are undiagnosed patients included in the analysis as the Bio*Me*™ Biobank. For example, an individual may not have a EHR record of AN in the Bio*Me*™ Biobank if (s)he had an AN-related visit at another healthcare facility or when they were adolescents^7^ (the youngest individual included in this study is 25 years old). Given the low prevalence of AN (∼1%)^41^ and our exclusion of all individuals with any ED diagnosis, we expect the impact of diagnostic contamination should be minimal. In addition, we note that our analysis relies on PRS derived from a European AN GWAS^22^, reducing predictive accuracy in the African-American and Hispanic individuals in our biobank. In order to increase power, we selected 5% weight change as the primary cutoff for weight trajectory phenotypes, given there are more individuals with weight changes at the 5% cutoff, compared to the 10% cutoff. Indeed, we only observed a significant association of AN-PRS with weight loss trajectory using the 5% cutoff, even though consistent effect direction was observed regardless of the cutoffs.

Despite previous evidence for sex-specific effects of AN-PRS on adolescent weight trajectories^25^, we did not see any sex-specific effects (P_interaction_=0.58), perhaps reflecting differing roles for AN genetics in weight trajectories throughout development. Similarly, despite evidence for a mitigation effect of obesity-PRS on weight loss in high AN-PRS individuals in the same study,^25^ we do not observe any mitigation effect of obesity-PRS on AN-PRS (i.e., top vs bottom decile AN-PRS OR on weight loss changed from 1.95 to 1.94 after adjustment for obesity-PRS). A few factors may explain these inconsistent findings. First, our study compared weight loss to stable weight, while Abdulkadir et al. compared weight loss to weight gain,^25^ which we might expect to yield larger effect sizes. We also note differences in study population (ages 10-24, vs. ages 25-85 in our study), weight trajectory definitions (e.g., a priori vs. statistical weight trajectory derivations^25,29,42^), as well as the exclusion of patients with eating disorders from our study.

Our study relies on EHR data, yielding significant power. The extensive weight measurements available through the Bio*Me*™ Biobank made it possible to construct weight trajectories in a large sample size of 21,487, spanning nine years on average. However, EHR-based analyses also have notable limitations. First, since weight measures are derived from existing records, rather than collected specifically for this study, we cannot distinguish unintentional from intentional weight loss. Second, weight measures are taken on an ad-hoc basis, with no set time frame, and measurement error may occur. However, one past study has compared clinic-measured weight against researcher-measured weight and showed a very high correlation between the two weight measures (correlation=0.99).^43^ In our study, the weight trajectory phenotypes achieved an accuracy of 98% or higher (**S. Table 1**) by minimizing the misclassification of weight measures in two ways: 1) by studying the overall weight trajectory using annual weights, and 2) by removing implausible weight outliers. Nevertheless, it is worth pointing out that even though we reduced error in weight measures via studying the annual weights, we lost the resolution to model any weight change happening within a year. Third, although we excluded participants with eating disorder or depression diagnoses, the participants in our study are patients in a large hospital system. Since only individuals with at least 3 annual weights in the Bio*Me*™ Biobank were included in this study, they could be less healthy in general and take various medications that can influence weight, compared to those who are healthy and rarely visit the doctors. Thus, findings in our study may not generalize to healthy populations.

To summarize, maintaining a stable weight is an indicator of well-being, demonstrated by its negative associations with various diseases, while fluctuating weight is associated with diseases such as depression. In addition, we find that adult weight trajectories are heritable and have genetic links with AN, such that individuals with higher genetic risk of developing AN are more likely to lose weight in adulthood, independent of the disorder itself. Future studies are necessary to identify shared and distinct genetic pathways of AN and obesity related to weight regulation.

## Supplementary Materials

### Supplementary Methods

#### Study Population

Participant enrollment in the Bio*Me*™ Biobank started in 2007 and occurred through clinical care sites when the patients visited their healthcare facilities/providers in the Mount Sinai Health System, which has a broad coverage of healthcare service in and around the New York City area.^44^ During enrollment, patients were interviewed in a systematic manner based on a questionnaire that collected demographic, lifestyle, and family medical history information and underwent a blood draw.^45^

#### Genotyping

SHAPEIT/IMPUTE2 was used for imputation with the 1000 Genomes Phase 3 as the reference panel.^28^ We QCed genotype data to include only unique SNPs with imputation INFO score > 0.7. The QC filters applied to retain the SNPs and individuals were: genotype missingness < 0.02, Hardy-Weinberg Equilibrium P > 10^−6^, MAF > 0.0001, individual missingness < 0.02, autosomal heterozygosity F_het_ value within 3 SD, family relatedness < 0.125 (one individual of each pair of 1^st^ or 2^nd^ degree relatives was removed).

#### Weight Trajectory

Extensive data cleaning was performed for the longitudinal weight measures, including (1) excluding weight outliers (> 4.5 SD of the mean weight per individual, or the ratio of maximum/minimum weight > 2.5 per individual); (2) removing duplicated weight measures taken on the same day (if more than one weight value was entered on the same day, the weight closest to the average weight in that year [if other weight records exist in that year] or the mean weight across all measures was retained); (3) removing observations with impossible weight change by manually inspecting the two ends of weight change distribution. For example, one weight measure for an individual was removed manually (+140kg in ten months, followed by −115kg in the following 14 months). Such ‘impossible’ weight gain/loss patterns more likely reflect clinician or data entry errors, conversion errors between weight units, or insurance card sharing (i.e., weights stemming from different patients), rather than true biological weight change.

Weight trajectories are defined as follows:

##### Stable weight trajectory

Maximum weight change from first annual weight < 5% or 10%.

##### Weight gain trajectory

Defined according to three criteria: (1) a net weight gain of > 0 kg over the measured period; (2) the maximum weight gain from baseline was ≥ 5%; (3) the individual had more weight gain than weight loss over time, and this was quantified by (a) a maximum weight loss of < 5% from baseline, or (b) the amount of maximum weight loss from baseline was < 45% of the overall weight change magnitude (maximum - minimum), so that the overall trend is still a weight gain. A cutoff of 45% was chosen based on manual visual inspection of the weight trajectory plots among a random group of 100 individuals. The same approach was taken when using a cutoff of 10% to define individuals with large weight gain.

##### Weight loss trajectory

Defined by: (1) a net weight loss of > 0 kg over the measured period; and (2) the maximum weight loss from baseline was ≥ 5%; and (3) the individual had more weight loss than weight gain over time, and this was quantified by (a) a maximum weight gain of < 5% from baseline, or (b) the amount of maximum weight gain from baseline was < 45% of the overall weight change magnitude (maximum - minimum), so that overall the trend is still a weight loss. The same approach was taken when using a cutoff of 10% to define individuals with large weight loss.

##### Weight cycle

Defined as a weight gain ≥ 5% from a previous weight at one interval and a weight loss ≥ 5% at another interval regardless of the chronological order. We used two approaches (local and global) to identify the cycles. First, for each individual, all of the weights were plotted in a chronological order. Inflection points (i.e., local highest and lowest weights) as well as the maximum and minimum weights of all measures (i.e., global highest and lowest weights) were identified. A *local* weight cycle is defined based on local highest and lowest annual weights (e.g., two local minimum weights and one local maximum weight can create a local weight cycle with the local maximum being the peak or the inflection point). Both the increase and decrease in weight on the two sides of the inflection point needs to be ≥ 5% (**S. Figure 9**). A *global* weight cycle was defined based on the global highest and lowest annual weights. A second highest and/or lowest annual weight was identified along with the global maximum and minimum annual weights to create a global weight cycle (**S. Figure 9**). The same approach was used when defining individuals with large weight cycles using 10% change as the cutoff. There were 72 participants (0.3%) who were weight cyclers under the 5% cutoff but were left as unclassified when using the 10% cutoff.

#### BioMe™ Phecodes

We required ≥ 2 instances of the same ICD-10 code for inclusion as a case for each phecode. Individuals with only one instance of each ICD-10 code were treated as missing. For example, participants with at least 2 records of the ICD-10 code, F50.00, were converted into a case for the AN phecode (305.21). Those with only 1 record of the F50.00 ICD-10 code was set as missing.

Exclusion criteria were applied when selecting controls for each phecode. For example, in order to be a control for either the AN, eating disorder, or depression phecode, individuals with any mental disorders were excluded to minimize the potential case contamination in the control population. Phecodes that were mapped to a “child” phecode (e.g., 305.21-AN) were also mapped to the “parent” phecode (305.2-eating disorder). In addition, for any sex-specific phecode, only individuals with the specified sex were included, and the rest was set to missing (e.g., only females were included for the phecode 306.1-mental disorders during/after pregnancy).

#### PRS Calculation for AN, Depression, and Obesity

The PRS was calculated as the effect of all genetic variants/single nucleotide polymorphism (a total of j SNPs) on the phenotype divided by the total number of alleles for that individual (*M*_*i*_), so as to average the genetic effect by the number of included alleles (equation 2).

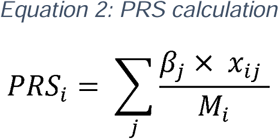

In this PRS formula, *i* is the *i* th individual.*j* is the *j*th SNP. β _j_ is the estimated effect size of the effect allele of each SNP on the phenotype of interest. *x*_ij_ is the exact genotype of the *j*th SNP in the *i* th individual (i.e., the number of effect alleles at each SNP position, could be 0, 1 or 2 assuming an additive genetic model). *β*_*j*_ could be generated through genome-wide association analysis or retrieved from previous GWAS, whereas the number of effect alleles of the *j*th SNP for the *i* th individual (*x*_*ij*_) requires individual genotype information.

Standard QC was performed for the PRS calculation, following the published PRSice protocol^46^. Ambiguous, duplicated, and mismatching SNPs were removed. PRS calculation and association with weight trajectories with adjustment of covariates were conducted using PRSice 2.2.2.^47^ Only SNPs that matched with those in the GWAS summary statistics were retained to calculate the corresponding PRS score (e.g., AN GWAS^22^ only included SNPs with MAF ≥ 0.01). For individuals with missing genotype of a certain SNP, a genotype value of 2 x MAF was assigned, based on the SNP MAF in the target data (Bio*Me*™). A best-fit p-value threshold for SNP inclusion in the PRS that explained the highest variation (R^2^) of the phenotype was chosen for each PRS-weight trajectory association.

#### Adjustment of Covariates

The disease/health covariates were created as binary variables based on ICD codes in the electronic health records. Key words were used first to search for people for each of the disease conditions, including cancer, COPD, HIV, AIDS, hypothyroid, hypothyroidism, end stage renal disease, ESRD, end stage renal failure, ESRF, dialysis, hemodialysis, and bariatric. The corresponding ICD codes with these diagnosis notes were then reviewed manually, and disease ICD code(s) or related ICD code(s) with potential influence on weight were adjusted. For example, individuals with a diagnosis note of either HIV or AIDS and a diagnosis code of HIV disease (i.e., B20 and B97.35 in ICD-10, 042 and 079.53 in ICD-9), or HIV-related complications (e.g., HIV complicating pregnancy/childbirth [O98.71, O98.72 in ICD-10], cachexia [R64 in ICD-10]) were included in the binary HIV covariate and adjusted in the model. Information about pregnancy was extracted from the obstetrics and gynaecology records and adjusted in the model using a pregnancy binary covariate (i.e., current, ectopic, gravida, para, preterm, term) except for abortion. Abortion was not included because this usually occurs in the first trimester when the increase in weight is minimal.

### Supplementary Results

#### PheWAS associations using 10% as the clinically relevant weight change cutoff

Previous clinical investigations of weight gain and loss have also considered 10%,^30,48,49^ in addition to 5%, as a clinically relevant weight change cut-off, therefore we repeated our analyses to construct weight trajectories with 10% threshold.

We observed moderate-to-high phenotypic correlations between weight trajectory defined using different cutoffs (5% or 10%) such that the within-group phenotypic correlations for the four weight trajectories ranged from 0.45 to 0.72 (e.g., correlation between weight loss 5% vs. weight loss 10% = 0.72, **S. Figure 2**).

Our main PheWAS findings remained significant when testing at the 10% threshold, including an association between weight cycle and depression (p=9.0×10^−10^), between weight gain and obesity phecodes (p ≤ 3.7×10^−49^), as well as between weight loss and protein-calorie malnutrition (p=1.9×10^−36^), gastrointestinal complications (p=5.1×10^−30^), bariatric surgery (p=1.6×10^−31^), and end stage renal disease (p=1.3×10^−36^) (**S. Figure 10, S. Table 6**).

Similarly, several sex-stratified PheWAS findings also remained significant using the 10% cutoff. For example, the sex difference in the weight loss association with anemia remained significant using the 10% cutoff (OR_male_=1.9, OR_female_=1.3, p_diff_=0.0004, **S. Table 8**). In addition, the sex-specific associations between weight cycle and swelling of limb and vitamin B complex deficiency were also significant at the 10% threshold (**S. Table 8**).

However, for the weight loss associations with osteoporosis (**S. Table 8**), we observed a greater effect magnitude in females using the 10% cutoff (OR_female_=1.35, p_female_=2.9×10^−5^, OR_male_=1.07, p_male_=0.70), though the sex difference was not significant (p_diff_=0.23).

In addition, we identified a sex-specific association between weight loss and dementia when the 10% cutoff was used. The dementia-weight loss association was much stronger in females (OR=3.0, p=5.8×10^−15^) compared to males (OR=1.8, p=0.0039, p_diff_=0.025), with similar findings found at the 5% cutoff (OR_female_=2.2, OR_male_=1.5) even though the sex difference was not significant at the 5% cutoff (p_diff_=0.14, **S. Table 7-8**). Similarly, we observed that males with edema were more likely to be associated with weight cycle (OR_male_=2.3 vs OR_female_=1.7, p_diff_=0.046, **S. Table 8, S. Figure 5**) and less likely to have a stable weight using the 10% weight cutoff (OR_male_=0.44 vs OR_female_=0.65, p_diff_=0.0058, **S. Table 8, S. Figure 11**), with similar sex difference seen using a 5% cutoff, though not all significant (**S. Table 7)**.

#### AN-PRS and eating disorder phecode association with weight trajectory

Little association was found for AN-PRS with the weight gain or weight cycle trajectories (empirical P ≥ 0.19).

To examine the AN-PRS association with weight loss trajectory on a phenotypic level, we tested the association between the eating disorder phecode (305.2) with weight loss trajectory (**Figure 2**). A consistently positive association was observed between the eating disorder phecode and the weight loss trajectory in the European ancestry group, though the confidence interval was wide due to a small number of cases (n=9). These 9 cases were females of European ancestry who have no missing covariates and have either a weight loss or stable weight trajectory, since the outcome of interest in this analysis is the weight loss trajectory with stable weight being the reference group.

#### Little association between AN-PRS and frequency of doctor visits

One question of interest was if the weight loss was intentional or not for those individuals with a weight loss trajectory. Since the EHR includes only objective weight measures, we hypothesized that weight loss among those with a higher AN-PRS was more likely to be intentional if they had fewer doctor visits (i.e., a proxy of overall well-being, and thus a weight loss trajectory is more likely to be intentional). Based on this, we examined the correlation of AN-PRS with number of encounter visits among individuals with a weight loss trajectory, but found that there was very little correlation (Pearson r = −0.003).

## Data Availability

Full PheWAS summary statistics and codes for constructing the longitudinal weight trajectories in a biobank setting are available on GitHub (https://github.com/xuj18/BioMe_weight_project).

## Data sharing

Full PheWAS summary statistics and codes for constructing the longitudinal weight trajectories in a biobank setting are available on GitHub: (https://github.com/xuj18/BioMe_weight_project).

## Acknowledgements

This work was supported in part through the computational resources and staff expertise provided by Scientific Computing at the Icahn School of Medicine at Mount Sinai under award number S10OD018522 from the Office of Research Infrastructure of the National Institutes of Health. Furthermore, this work was supported through the resources and staff expertise provided by the Charles Bronfman Institute for Personalized Medicine and The Bio*Me*™ Biobank Program at the Icahn School of Medicine at Mount Sinai. In addition, we would like to thank the Huckins Lab members for their feedback on this project during the lab meetings. The content is solely the responsibility of the authors and does not necessarily represent the official views of the National Institutes of Health.

## Contributors

JX and LMH conceptualized the study. JX created the method for weight trajectory classification, conducted the analysis, and drafted the manuscript. LMH advised on the development of the method and the analyses. JSJ advised on the code and verified the underlying weight trajectory data. AB, JJ, MAK, ML, SLM, NGM, PBM, LVP, LMT, CMB in the PGC-ED working group contributed to the data collection for the AN GWAS summary statistics. JX and LMH revised the manuscript. All authors were involved in reviewing the manuscript before submission.

