## Supplemental figures for "Exploring the clinical consequences and genetic aetiology of adult weight trajectories"

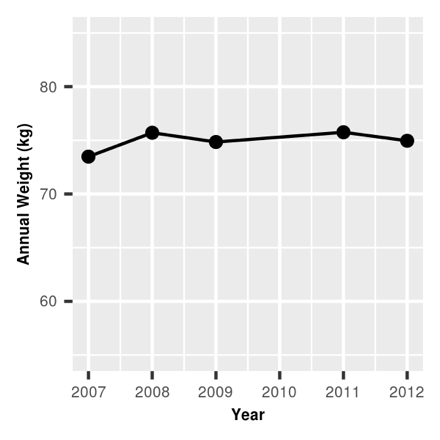

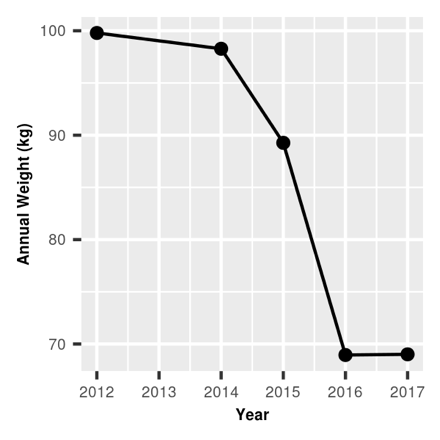

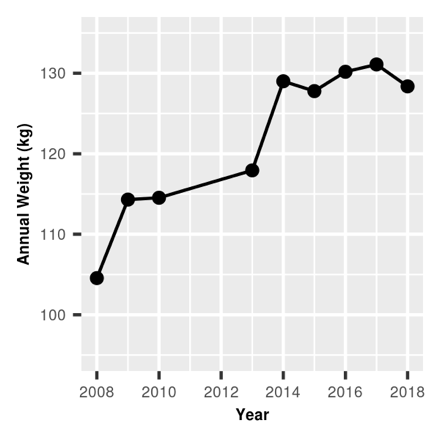

**C**

**B**

**A**

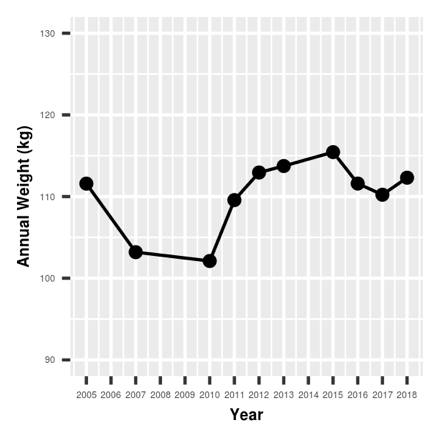

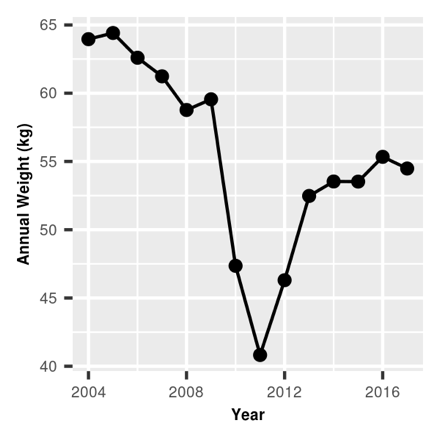

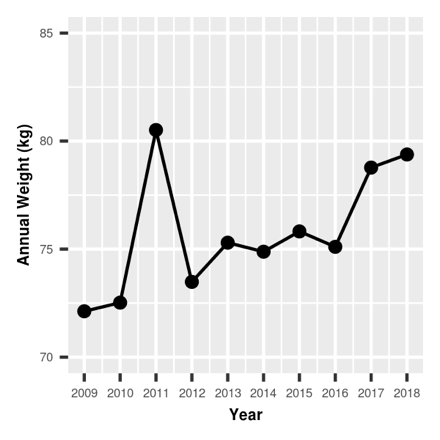

**E**

**D**

**F**

**Supplemental Figure 1. Examples of different types of individual weight trajectories based on annual weights.** (A) stable weight trajectory (B) weight loss trajectory (C) weight gain trajectory (D) weight cycle trajectory (E) weight cycle + weight loss trajectory (F) weight cycle + weight gain trajectory

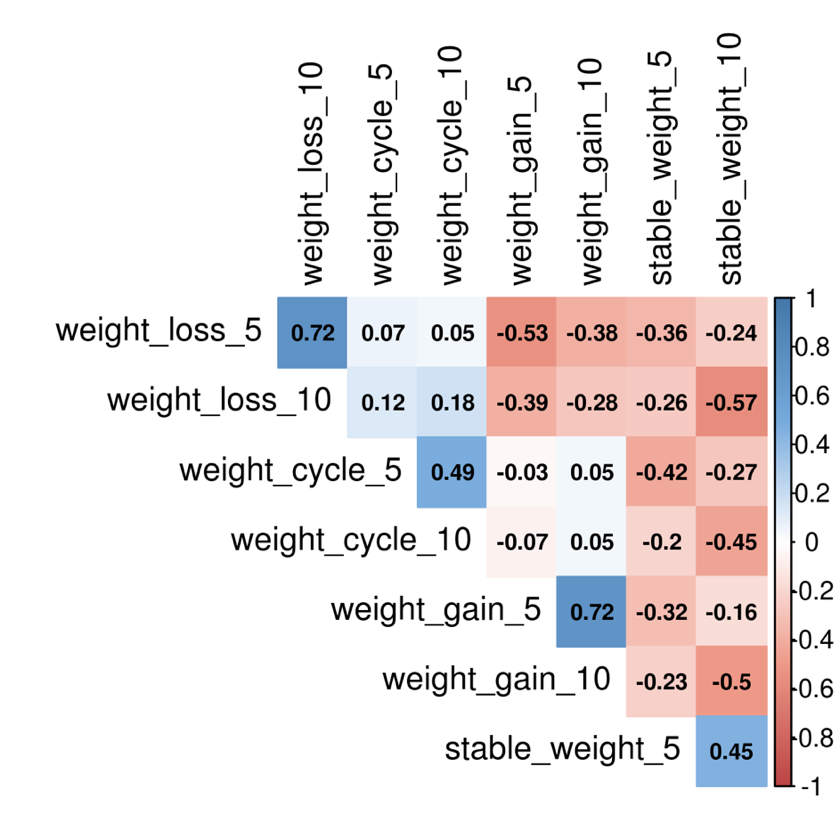

**A**

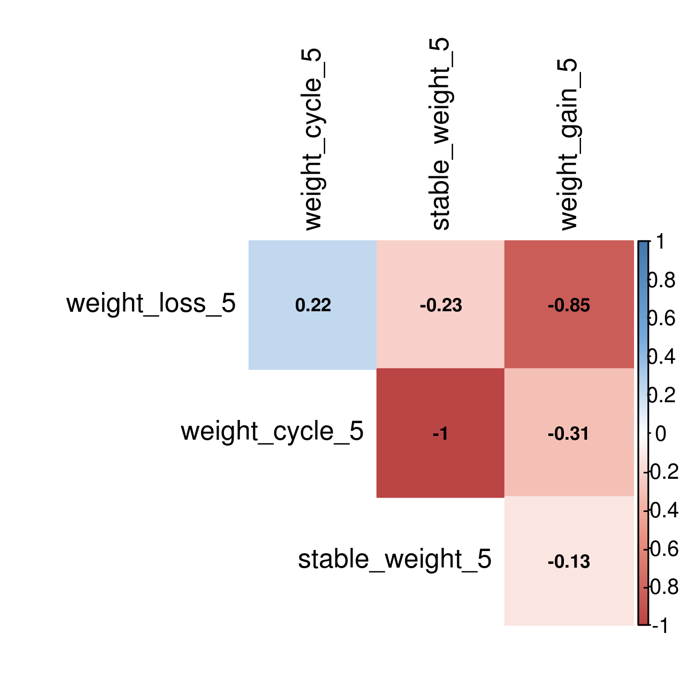

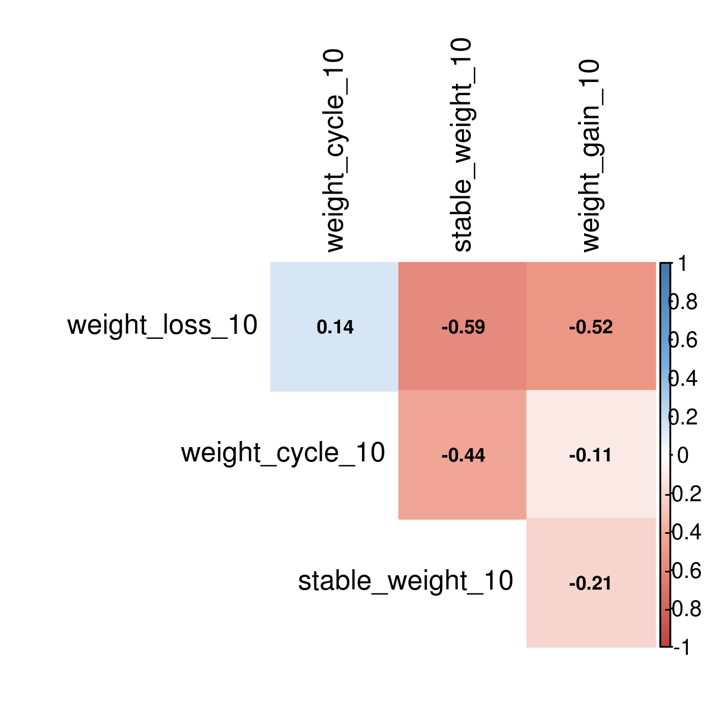

**C**

**B**

**Supplemental Figure 2. Correlation plot between four types of weight trajectories (stable weight, weight loss, weight gain, weight cycle). (A) phenotypic correlation between the different trajectories, (B) genetic correlation between the different trajectories using the 5% cutoff, (C) genetic correlation between the different trajectories using the 10% cutoff.** Two different weight change cutoffs (5% or 10%) were used for weight trajectory classification. The color intensity is proportional to the correlation coefficient shown. Positive correlation is displayed in blue while negative correlation is displayed in red. The baseline BMI status was adjusted for the genetic correlations between weight trajectories.

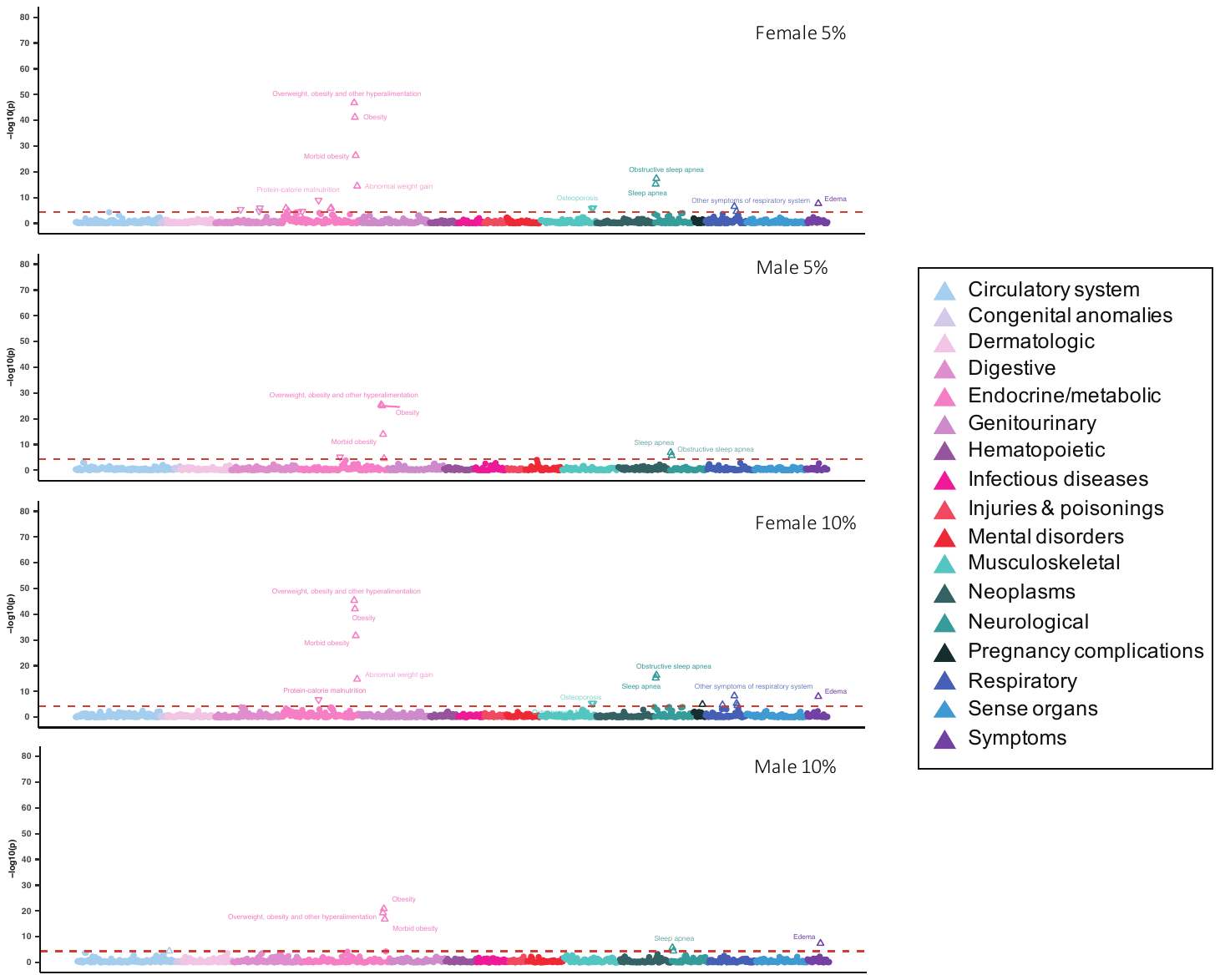

**D**

**C**

**B**

**A**

**Supplemental Figure 3. The phenome-wide plots of the associations of the weight gain trajectory with phecodes in the Bio*Me*^TM^ Biobank by sex under different weight change cutoffs (5%, 10%).** The red line denotes the Bonferroni-corrected p-value significance (P = 4.4$\times$10^-5^). All the significant phecodes above the red line are annotated. The phecodes are grouped into 17 different disease categories. An upward triangle (△) denotes a positive association, while a downward triangle (▽) denotes a negative association. (A) PheWAS plot for weight gain trajectory using 5% cutoff in females; (B) PheWAS plot for weight gain trajectory using 5% cutoff in males; (C) PheWAS plot for weight gain trajectory using 10% cutoff in females; (D) PheWAS plot for weight gain trajectory using 10% cutoff in males.

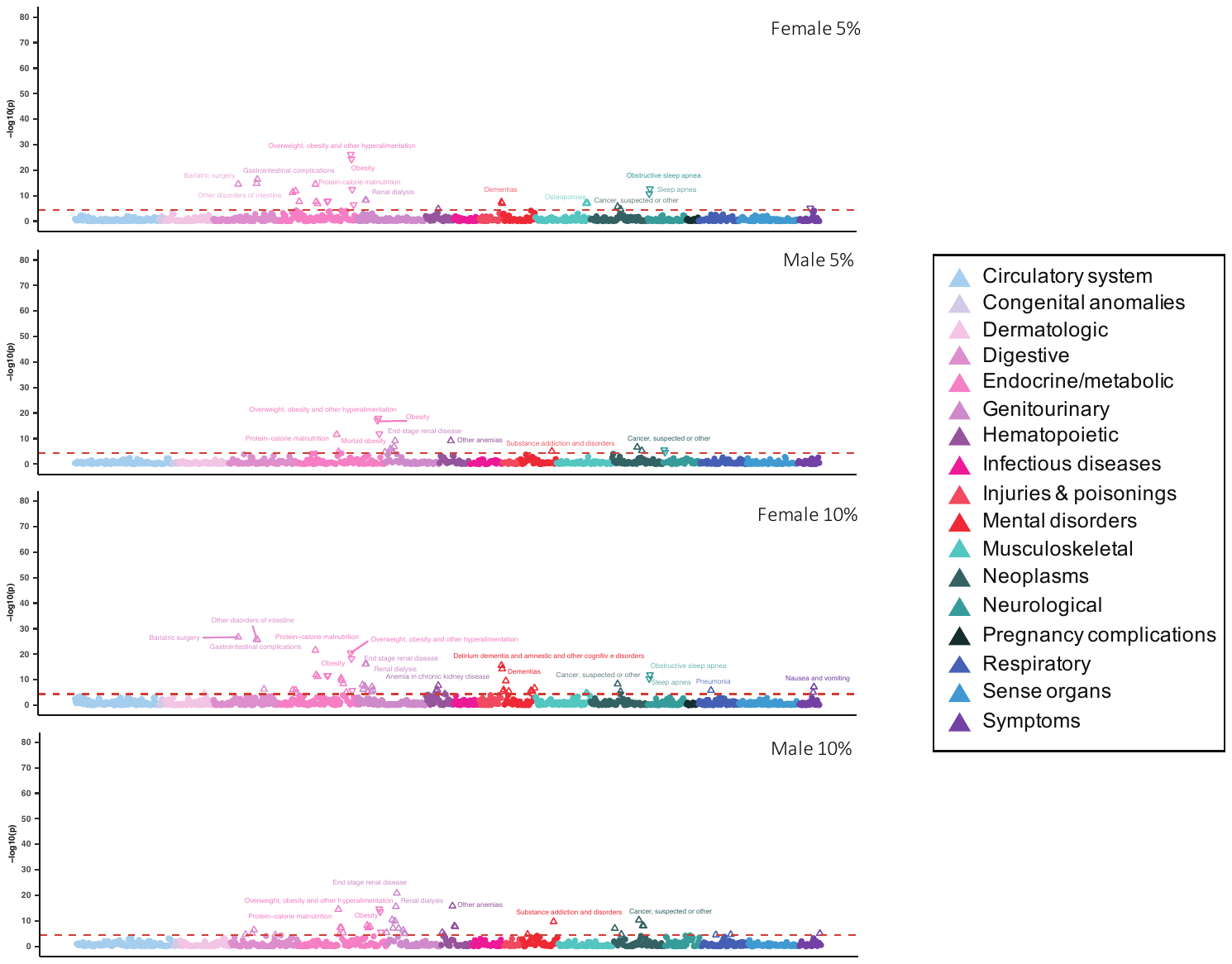

**D**

**C**

**A**

**B**

**Supplemental Figure 4. The phenome-wide plots of the associations of the weight loss trajectory with phecodes in the Bio*Me*^TM^ Biobank by sex under different weight change cutoffs (5%, 10%).** The red line denotes the Bonferroni-corrected p-value significance (P = 4.4$\times$10^-5^). All the significant phecodes above the red line are annotated. The phecodes are grouped into 17 different disease categories. An upward triangle (△) denotes a positive association, while a downward triangle (▽) denotes a negative association. (A) PheWAS plot for weight loss trajectory using 5% cutoff in females; (B) PheWAS plot for weight loss trajectory using 5% cutoff in males;

(C) PheWAS plot for weight loss trajectory using 10% cutoff in females; (D) PheWAS plot for weight loss trajectory using 10% cutoff in males.

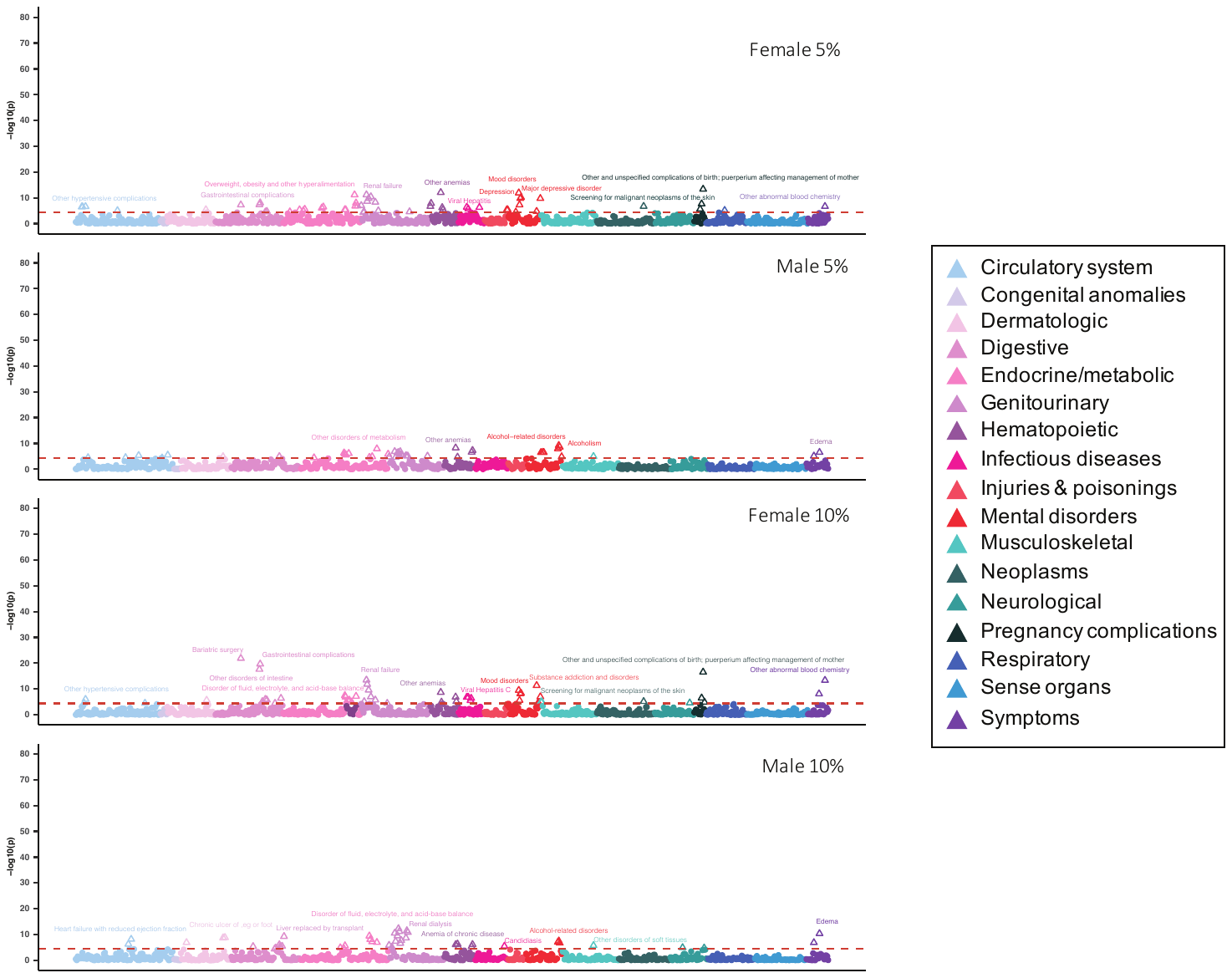

**D**

**C**

**B**

**A**

**Supplemental Figure 5. The phenome-wide plots of the associations of the weight cycle trajectory with phecodes in the Bio*Me*^TM^ Biobank by sex under different weight change cutoffs (5%, 10%).** The red line denotes the Bonferroni-corrected p-value significance (P = 4.4$\times$10^-5^). All the significant phecodes above the red line are annotated. The phecodes are grouped into 17 different disease categories. An upward triangle (△) denotes a positive association, while a downward triangle (▽) denotes a negative association. (A) PheWAS plot for weight cycle trajectory using 5% cutoff in females; (B) PheWAS plot for weight cycle trajectory using 5% cutoff in males;

(C) PheWAS plot for weight cycle trajectory using 10% cutoff in females; (D) PheWAS plot for weight cycle trajectory using 10% cutoff in males.

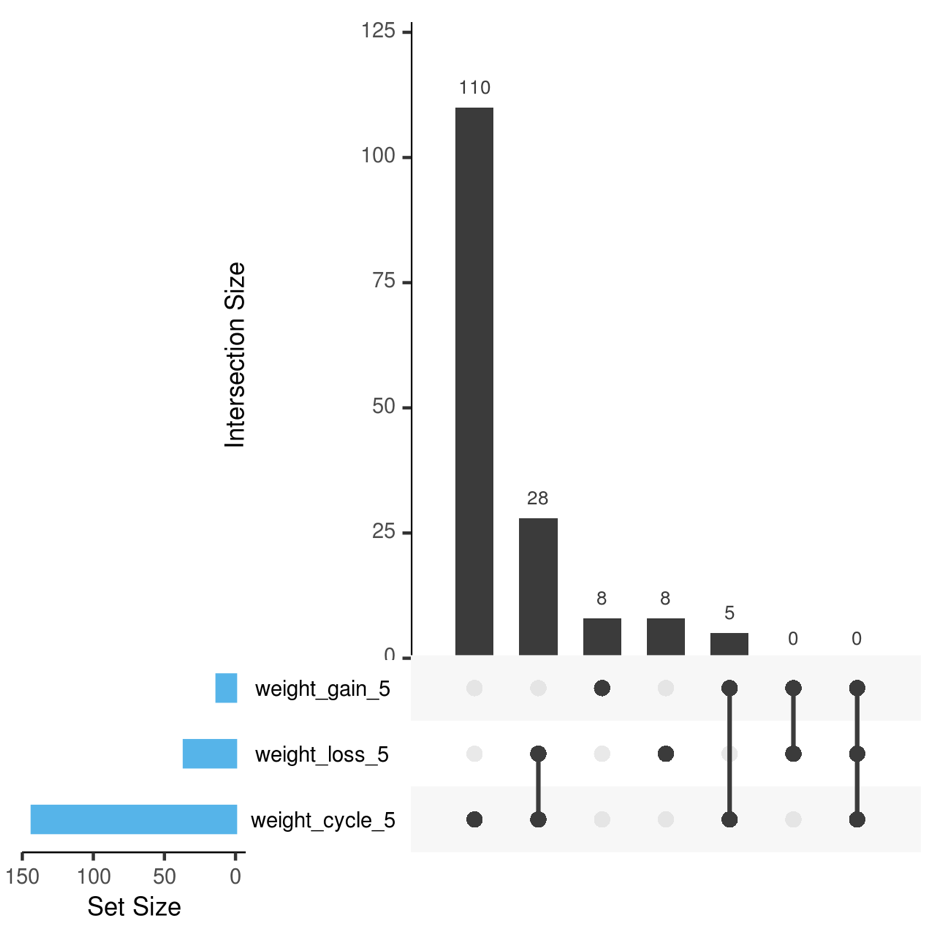

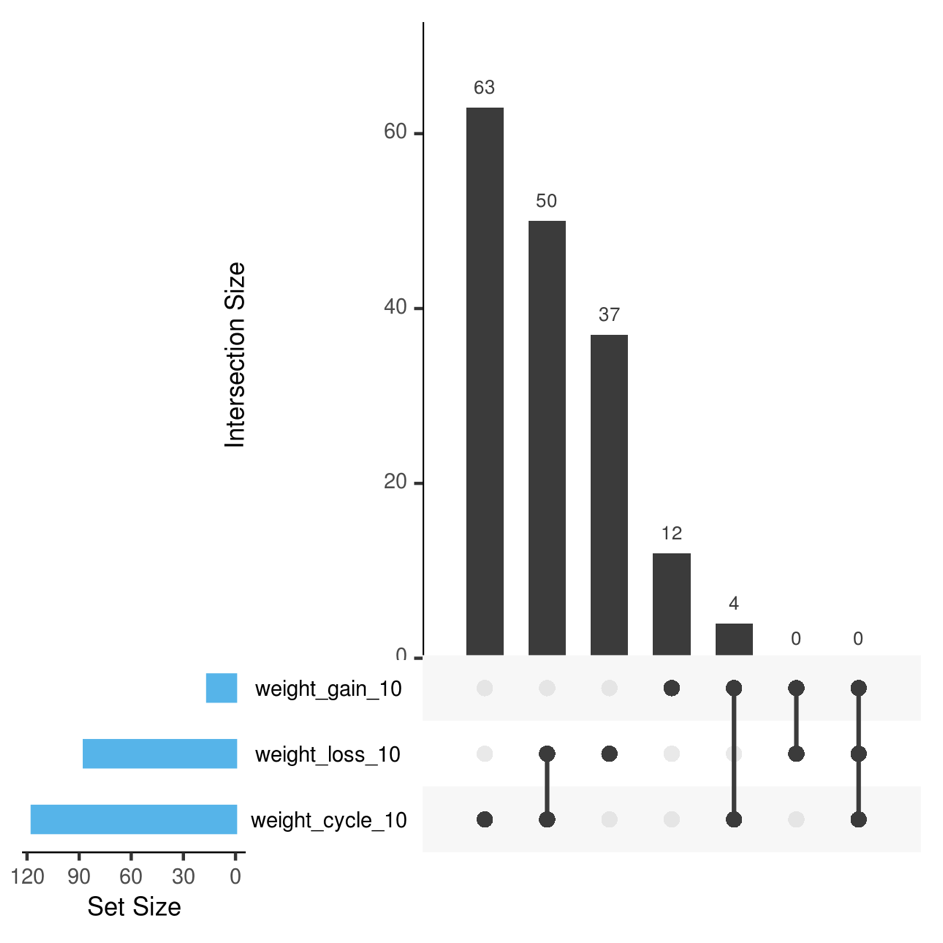

**B**

**A**

**Supplemental Figure 6. UpSet plot of distinct and shared disease phecodes associated with weight gain, weight loss, and/or weight cycle trajectories.** Only those phecodes that had significantly positive associations with each weight trajectory were included, with Bonferroni-corrected p-value $\leq$ 4.4$\times$10^-5^. Panel (A) shows the number of overlapped and unique phecodes falling in different categories when the weight trajectory was defined using the 5% weight change cutoff, whereas panel (B) shows the number of overlapped and unique phecodes falling in different categories when the weight trajectory was defined using the 10% weight change cutoff.

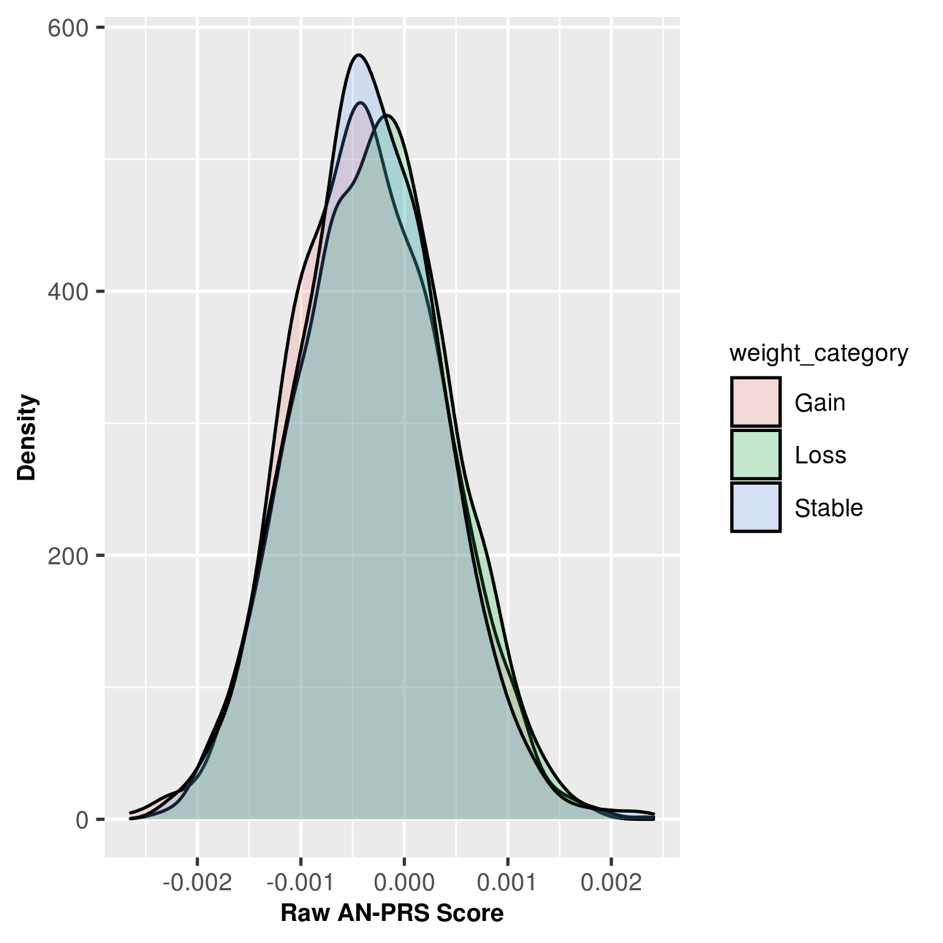

**Supplemental Figure 7. Distribution of raw AN PRS by three weight trajectory groups (weight gain, weight loss and stable weight).** The density score on the y-axis is proportional to the number of participants across the PRS score distribution. Abbreviation: AN, anorexia nervosa; PRS, polygenic risk score.

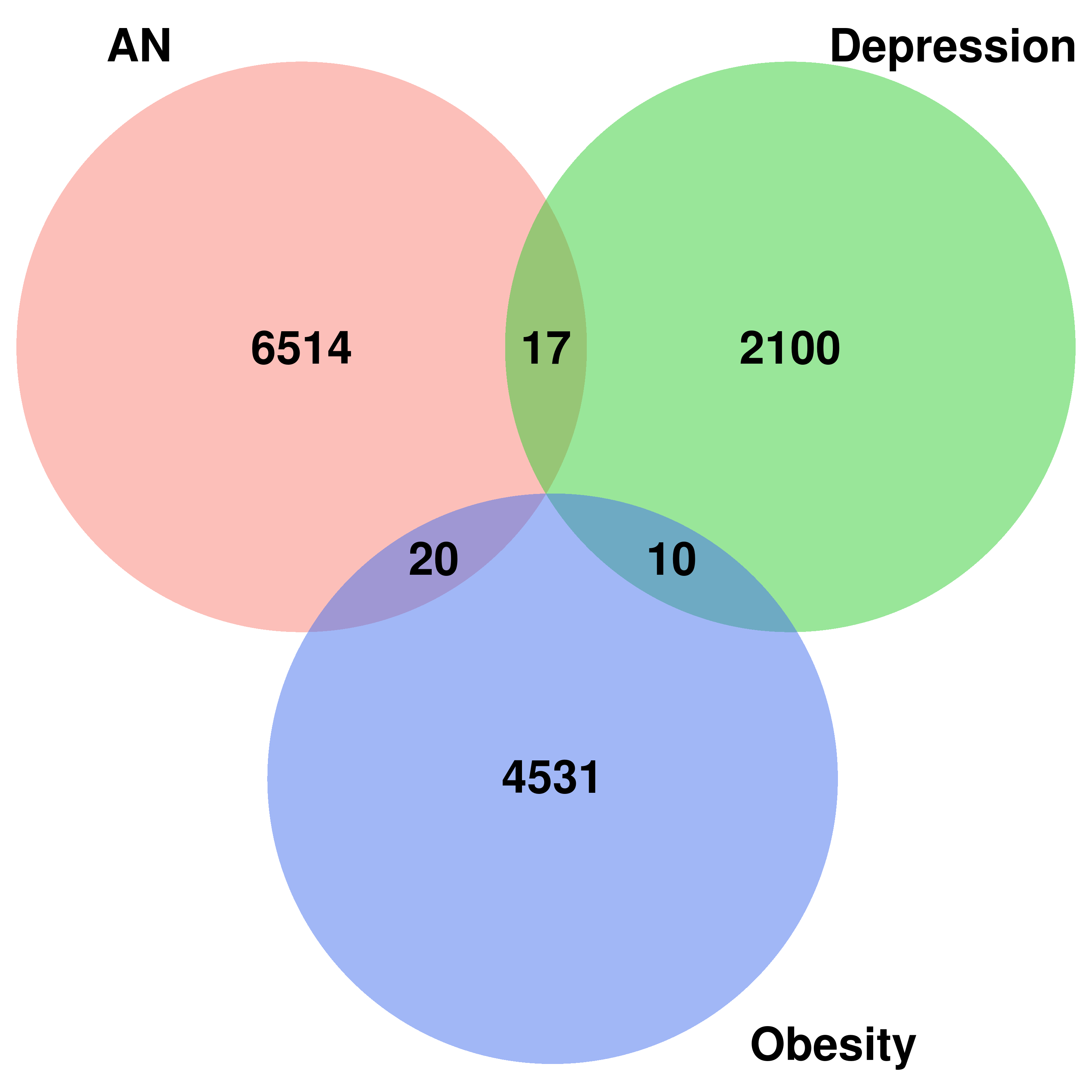

**Supplemental Figure 8. SNP overlap between three PRS scores.** This Venn diagram shows the overlap of genetic variant/SNPs that were included in each of the polygenic risk score (PRS) calculation for the analysis of genetic risk of anorexia nervosa (AN), depression, and obesity with weight loss trajectory based on the 5% weight change cutoff. For example, there were 17 SNPs included in the calculation of both AN and depression PRS, whereas there were 20 SNPs included in the calculation of both AN and obesity PRS calculation.

**B**

**A**

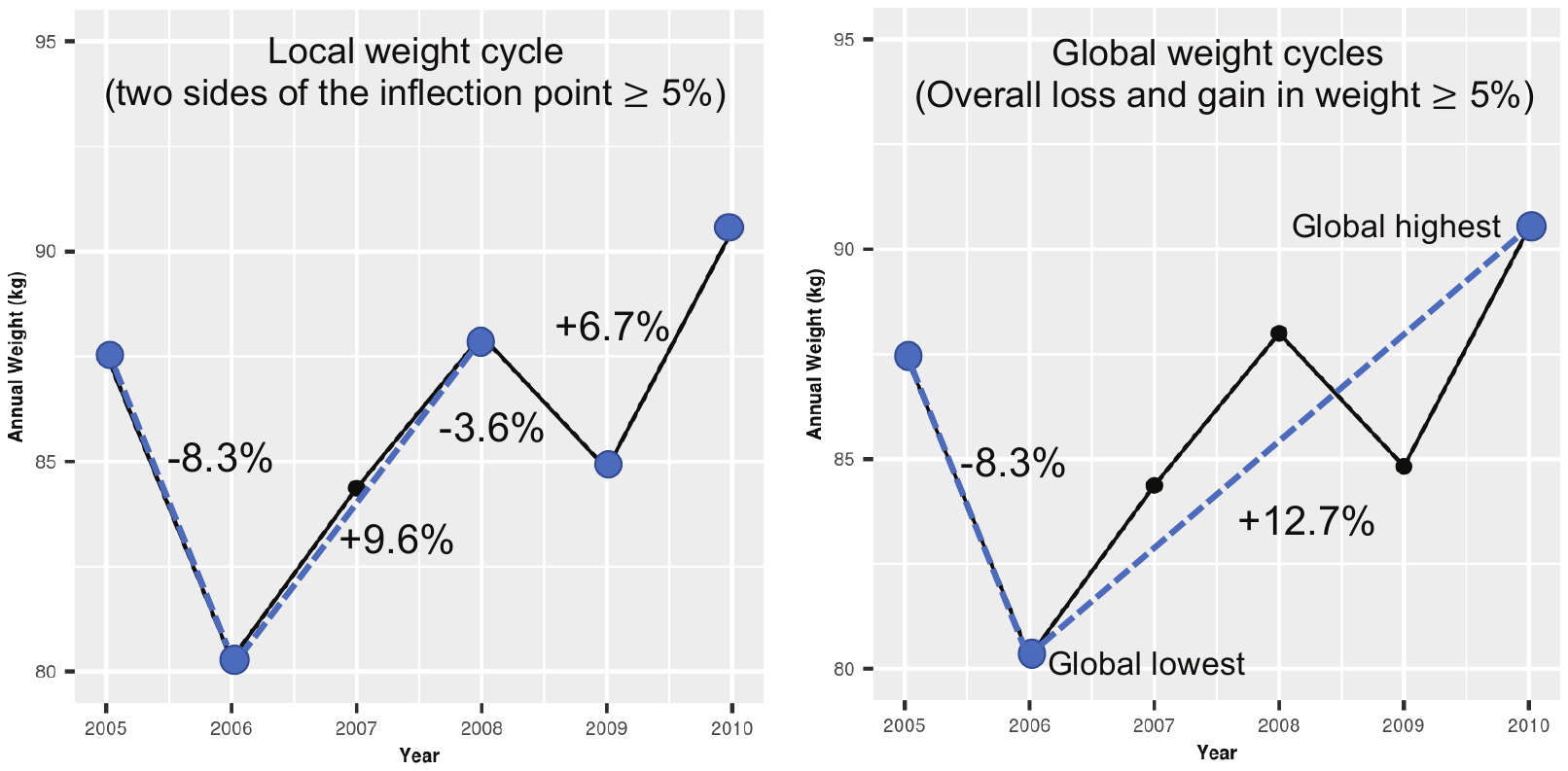

**Supplemental Figure 9. Definition of a local weight cycle (A) and a global weight cycle (B).** The local weight cycle was based on the percent of weight change between local highest and lowest weights. In panel (A), there is one local weight cycle from 2005 to 2008 as there was an 8.3% decrease in weight from 2005 to 2006, and a subsequent increase in weight from 2006 to 2008, with both of the absolute weight changes $\geq$ 5%. In panel (B), a global weight cycle was identified based on the percent of weight change from baseline to the global lowest weight (i.e., minimum weight for this individual, -8.3%) and from the global lowest weight to the global highest weight (i.e., maximum weight for this individual, +12.7%), with both of the absolute weight changes $\geq$ 5%. One individual will be counted towards having a weight cycle if he/she has a local and/or global weight cycle.

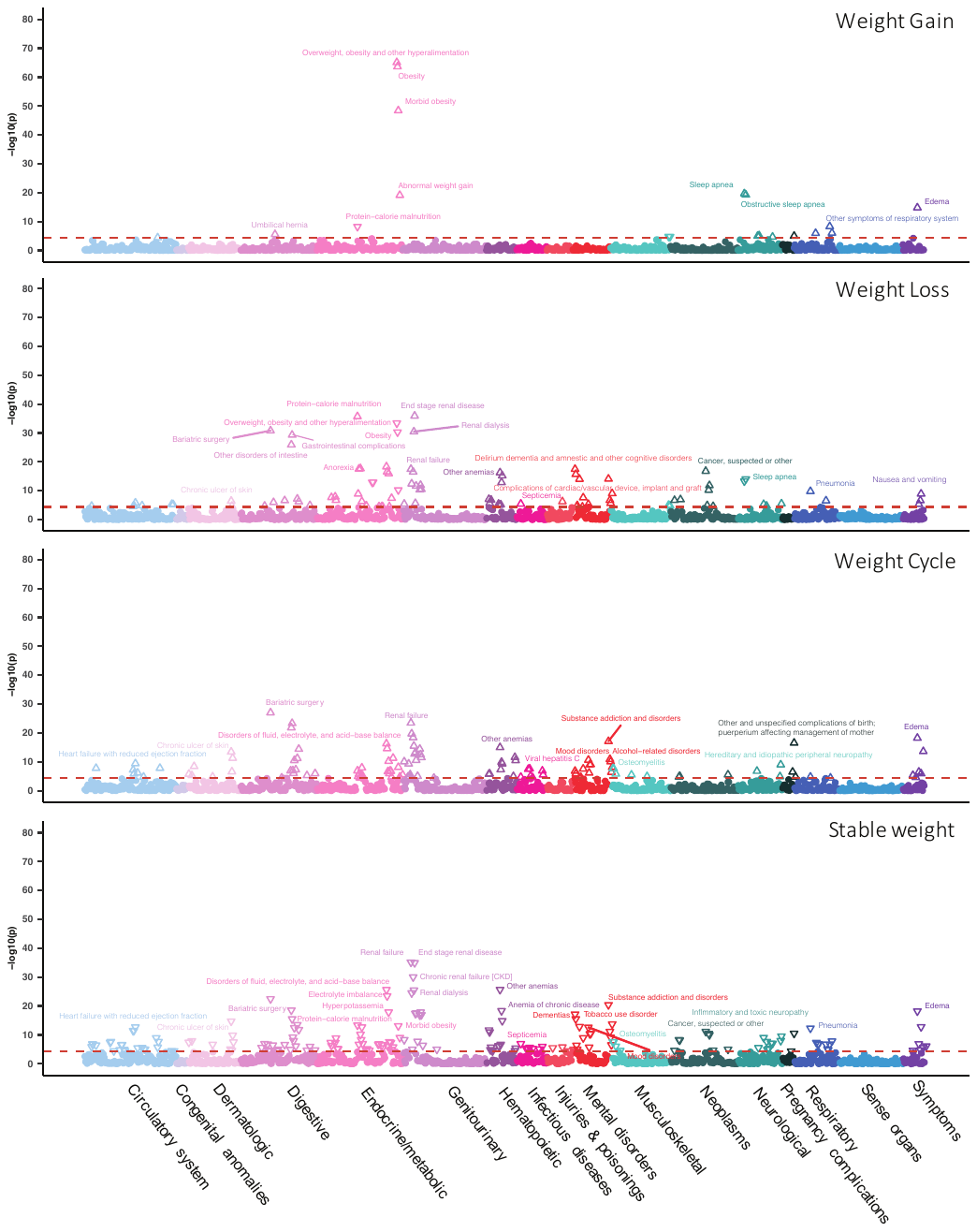

**D**

**C**

**B**
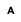

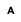

**A**

**Supplemental Figure 10. The phenome-wide plots of the associations of the weight trajectory with phecodes in the Bio*Me*^TM^ Biobank using the 10% weight change cutoff.** Phecodes above the blue line passed the Bonferroni-corrected p-value threshold (P < 4.4$\times$10^-5^). The phecodes are grouped into 17 different disease categories. An upward triangle (△) denotes a positive association, while a downward triangle (▽) denotes a negative association. The top associations are annotated in each plot. (A) PheWAS plot for weight gain trajectory; (B) PheWAS plot for weight loss trajectory; (C) PheWAS plot for weight cycle trajectory; (D) PheWAS plot for stable weight trajectory.

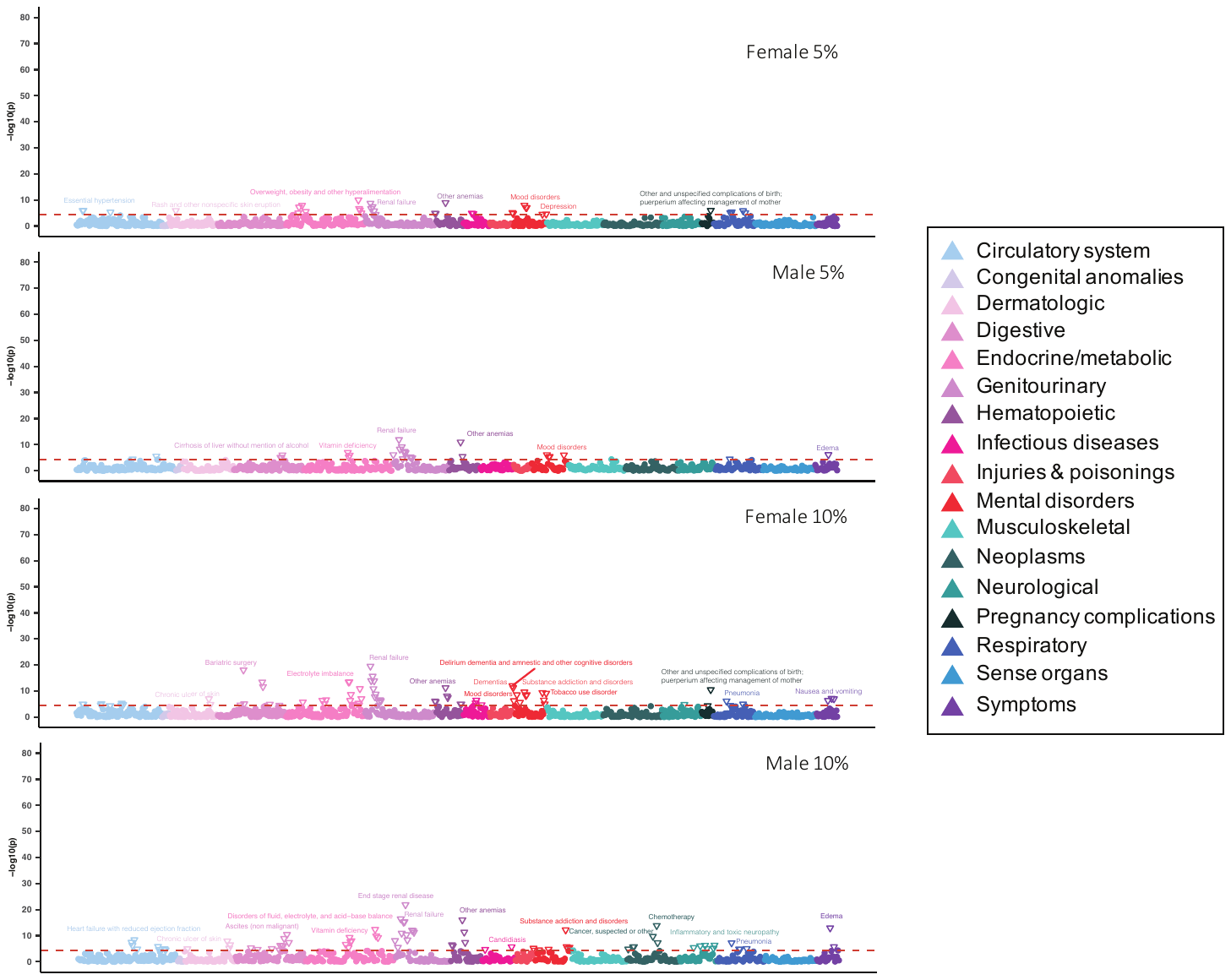

**D**

**C**

**B**

**A**

**Supplemental Figure 11. The phenome-wide plots of the associations of the stable weight trajectory with phecodes in the Bio*Me*^TM^ Biobank by sex under different weight change cutoffs (5%, 10%).** The red line denotes the Bonferroni-corrected p-value significance (P = 4.4$\times$10^-5^). All the significant phecodes above the red line are annotated. The phecodes are grouped into 17 different disease categories. An upward triangle (△) denotes a positive association, while a downward triangle (▽) denotes a negative association. (A) PheWAS plot for stable weight trajectory using 5% cutoff in females; (B) PheWAS plot for stable weight trajectory using 5% cutoff in males; (C) PheWAS plot for stable weight trajectory using 10% cutoff in females; (D) PheWAS plot for stable weight trajectory using 10% cutoff in males.
